# Genetic determinants of blood gene expression and splicing and their contribution to molecular phenotypes and health outcomes

**DOI:** 10.1101/2023.11.25.23299014

**Authors:** Alex Tokolyi, Elodie Persyn, Artika P. Nath, Katie L. Burnham, Jonathan Marten, Thomas Vanderstichele, Manuel Tardaguila, David Stacey, Ben Farr, Vivek Iyer, Xilin Jiang, Samuel A. Lambert, Guillaume Noell, Michael A. Quail, Diana Rajan, Scott C. Ritchie, Benjamin B. Sun, Scott A.J. Thurston, Yu Xu, Christopher D. Whelan, Heiko Runz, Slavé Petrovski, Daniel J. Gaffney, David J. Roberts, Emanuele Di Angelantonio, James E. Peters, Nicole Soranzo, John Danesh, Adam S. Butterworth, Michael Inouye, Emma E. Davenport, Dirk S. Paul

**Author notes:** Correspondence: Dirk Paul. These authors contributed equally (Alex Tokolyi, Elodie Persyn, Michael Inouye, Emma E. Davenport, Dirk S. Paul).

## Abstract

The biological mechanisms through which most non-protein-coding genetic variants affect disease risk are unknown. To investigate the gene-regulatory cascades that ensue from these variants, we mapped blood gene expression and splicing quantitative trait loci (QTLs) through bulk RNA-sequencing in 4,732 participants, and integrated these data with protein, metabolite and lipid QTLs in the same individuals. We identified *cis*-QTLs for the expression of 17,233 genes and 29,514 splicing events (in 6,853 genes). Using colocalization analysis, we identified 3,430 proteomic and metabolomic traits with a shared association signal with either gene expression or splicing. We quantified the relative contribution of the genetic effects at loci with shared etiology through statistical mediation, observing 222 molecular phenotypes significantly mediated by gene expression or splicing. We uncovered gene-regulatory mechanisms at GWAS disease loci with therapeutic implications, such as *WARS1* in hypertension, *IL7R* in dermatitis and *IFNAR2* in COVID-19. Our study provides an open-access and interactive resource of the shared genetic etiology across transcriptional phenotypes, molecular traits and health outcomes in humans (https://IntervalRNA.org.uk).

## Introduction

The majority of genetic variants associated with common diseases and other complex traits identified through genome-wide association studies (GWAS) lie in non-protein-coding sequences.^1^ Consequently, the molecular mechanisms that underpin many of these genotype–phenotype associations are unclear. Molecular quantitative trait locus (QTL) mapping studies, which identify genetic determinants of transcript, protein or metabolite abundance, can address this knowledge gap by identifying the molecular intermediaries that mediate genetically driven disease risk. These studies can provide specific hypotheses for functional validation experiments.^2,3^

Molecular QTL data can be used for a range of biomedical applications. For example, they have the potential to identify and validate new therapeutic targets and pathways; inform about the biological mechanisms of drug action and safety; highlight novel therapeutic indications; and reveal clinically relevant biomarkers.^4–6^

Many previous studies have carried out QTL mapping within a single molecular domain such as expression (eQTL) or protein (pQTL) analysis.^7–12^ However, QTL data from multiple -omic modalities are needed to fully understand the causal molecular chain of events from genetic variation to complex trait phenotypes.^13^ Moreover, the availability of multi-modal data in a single population sample enhances the interpretation and validity of causal inference analyses. For example, mediation analysis in a single cohort presents a strategy for identifying phenotypes that share a common genetic pathway and for quantifying the proportion of the total genetic effect on those phenotypes.^14^

Here, we use the INTERVAL study^15,16^, a bioresource of approximately 50,000 blood donors with extensive multi-dimensional ‘omics’ profiling, to identify gene expression and splicing QTLs based on peripheral blood RNA-sequencing (n=4,732 individuals). Then, we integrate the QTL data with additional molecular QTL data derived from the same study (Figure 1). These data include plasma protein levels measured through an antibody-based proximity extension assay (Olink Target panels, n=4,662–4,981 individuals)^17,18^ and an aptamer-based multiplex protein assay (SomaScan v3, n=3,301)^5^, as well as serum metabolite levels measured using an untargeted mass spectrometry platform (Metabolon HD4, n=14,296)^10^ and a nuclear magnetic resonance spectroscopy platform (Nightingale Health, n=40,849)^19,20^.

**Figure 1.**
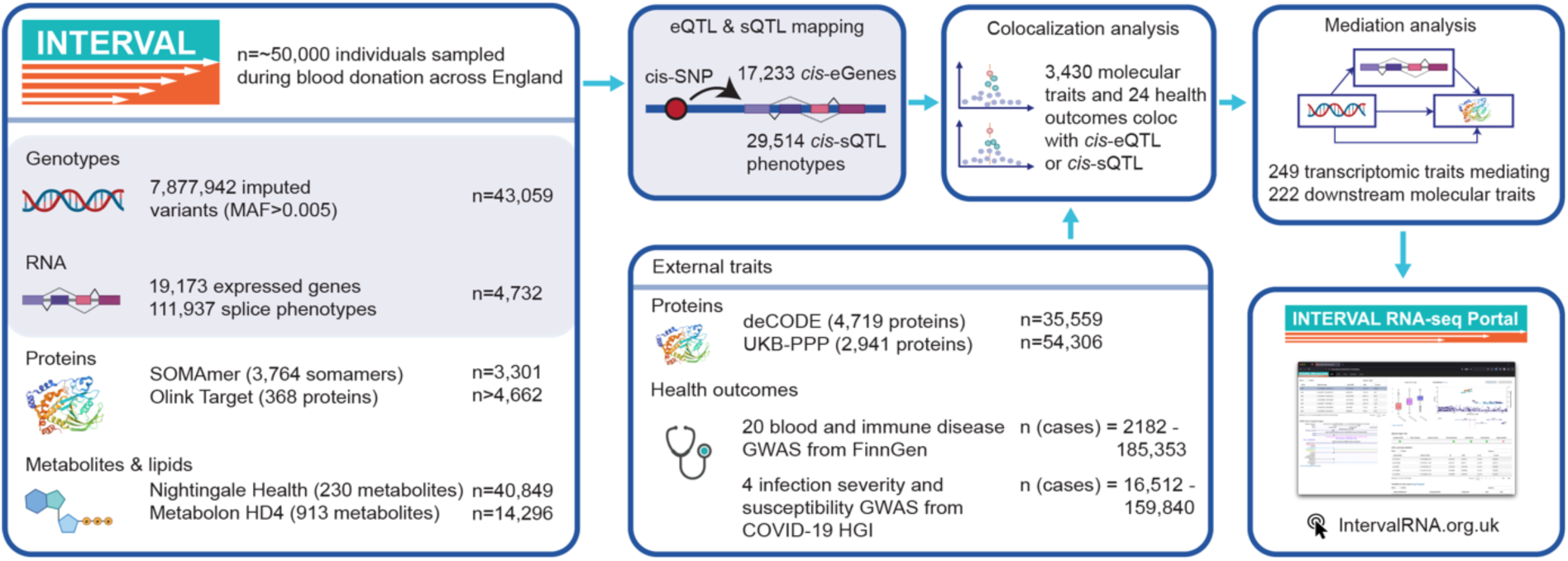
Overview of the multi-omic data available in the INTERVAL study and external cohorts, as well as the main analytical approaches. Abbreviations: UKB-PPP, UK Biobank Pharma Proteomics Project; COVID-19 HGI, COVID-19 Host Genetics Initiative; MAF, minor allele frequency; QTL, quantitative trait locus.

Our data reveal genetic effects on the expression and splicing of local and distant genes. We assess shared genetic etiology across molecular traits and health outcomes using statistical colocalization. Then, we further investigate the genetic effects on downstream molecular phenotypes through transcriptional events by conducting mediation analyses. Based on these analyses, we develop an open-access portal that enables exploration of this compendium of molecular QTLs (https://IntervalRNA.org.uk).

## Results

### Genetic regulation of local gene expression and splicing

We performed bulk RNA-sequencing on peripheral blood collected from 4,732 blood donors recruited as part of the INTERVAL study (Methods). The expression levels of 19,173 autosomal genes and 111,937 *de novo* transcript splicing phenotypes (herein referred to as “splicing events”) from differential intron usage ratios in 11,016 genes were quantified. Then, we mapped local (*cis*) expression QTLs (eQTLs) within ±1Mb of the transcription start site (TSS) and splicing QTLs (sQTLs) within ±500kb of the center of the spliced region.

We identified 17,233 genes (89.9% of the 19,166 tested) with at least one significant *cis*-eQTL (“*cis*-eGene”) at a false-discovery rate (FDR) <0.05 (Tables S1 and S2; Methods). To identify independent signals at each *cis*-eQTL, we performed stepwise conditional analyses (Methods). Across the transcriptome, this revealed 56,959 independent signals (53,457 unique lead variants), with a median of 3 independent signals per gene (range: 1–23; Tables S1 and S3). We compared our results to those from the eQTLGen Consortium study by Võsa *et al*, a meta-analysis of eQTL studies based on microarray and RNA-sequencing data (n=31,684 individuals).^9^ Z-scores from eQTL lead SNPs were highly correlated between these studies (Pearson r^2^=0.9; Figure S1; Tables S2 and S4). These results highlight the consistency of eQTL discovery results across independent datasets and mapping technologies.

Next, we investigated genetic associations with splicing events, identifying 29,514 splicing event phenotypes with a *cis*-sQTL at FDR<0.05 (Tables S1 and S5). These splicing events with a *cis*-sQTL were mapped to 6,853 genes (“*cis*-sGenes”) with a median of 3 splicing events observed per *cis*-sGene (range: 1–128). This included 543 *cis*-sGenes that were not identified as *cis*-eGenes. The long non-coding RNA *FAM157C*, which is involved in cell proliferation and induction of apoptosis,^21^ contained the most splicing events (n=128, within 11 clusters defined by shared splice donor or acceptor sites). While this gene is known to contain 33 exons, the splicing events were mostly intronic (n=105/128) and rarely overlapping previously defined exon boundaries (n=9/128). Across all splicing events with *cis*-sQTLs, these had a median length of 1,549bp and excised a protein-coding sequence in 32.4% of cases (the remainder related to intronic and UTR excisions). The median distance from the *cis*-sQTL lead variants to the center of the splicing event was 187bp upstream, with lead variants forming a bimodal distribution around the start and end of the sGene (Figure 2A).

**Figure 2.**
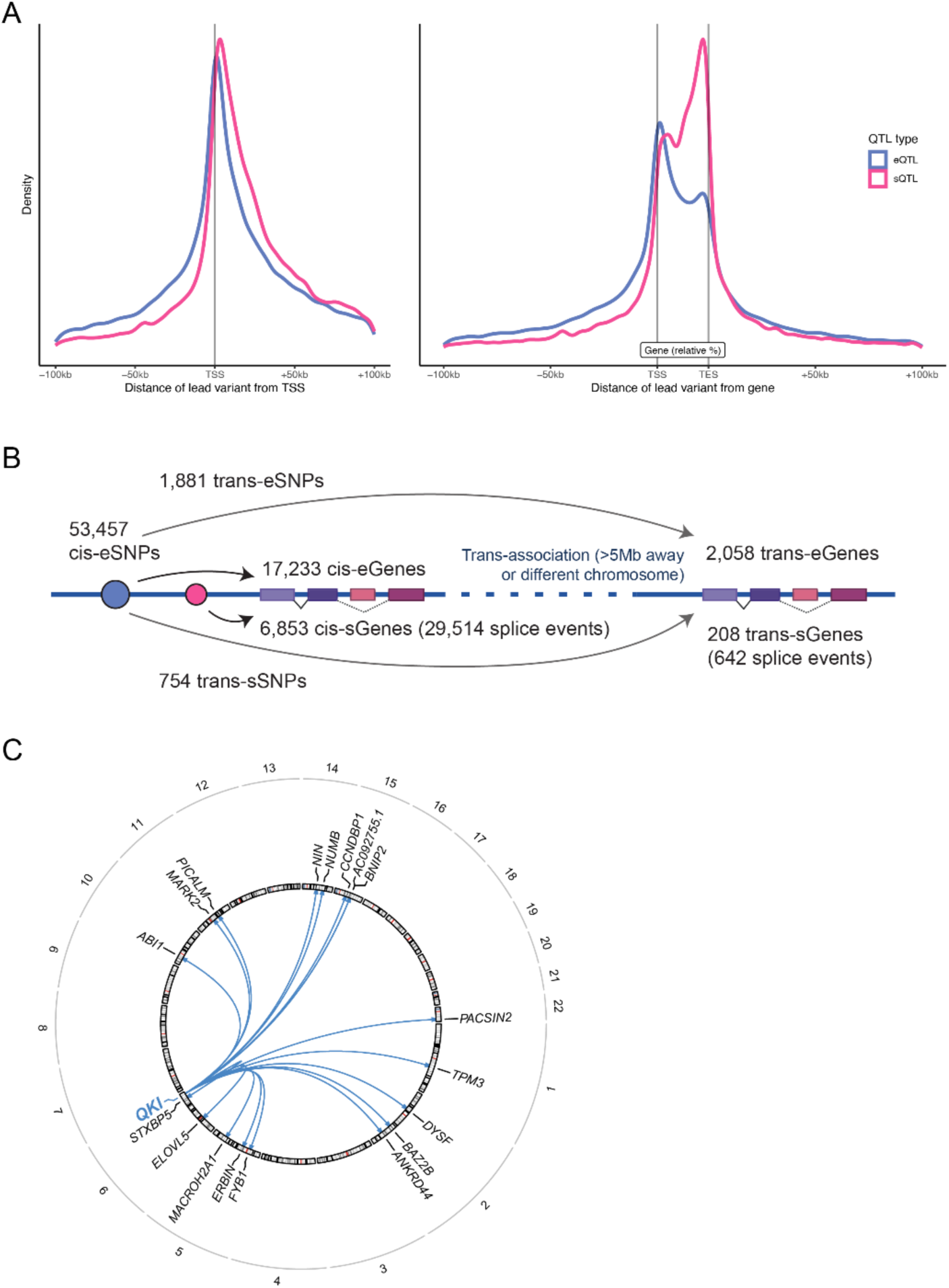
Genetic influences on gene expression and splicing. **A)** Distribution of lead variants at *cis*-eQTLs and *cis*-sQTLs around the TSS and gene body (normalized to the median gene length of 24kb). **B)** Schematic of the *trans*-QTL mapping analysis approach and summary of the QTL discovery results. **C)** Circos plot of the *trans*-splicing of 18 sGenes by the *cis*-eQTL for *QKI*. Abbreviations: TSS, transcription start site; TES, transcription end site.

After conditional analysis for each *cis*-sQTL, we identified 47,050 independent signals (34,205 unique lead variants), with a median of one independent signal per *cis*-sQTL (range: 1–20; Tables S1 and S6). To characterize independent variant effects on transcript splicing, we compared primary and secondary *cis*-sQTLs. Primary *cis*-sQTL signals were enriched within the gene body of sGenes compared to secondary signals (p=2.84×10^-314^, chi-squared test; Figures 2A and S2). Primary *cis*-sQTL signals were more enriched towards the transcription end site (median of 17.36kb downstream of the TSS) compared to *cis*-eQTLs with a median of 5.51kb downstream of the TSS (p=8.42×10^-259^, Wilcoxon test; Figure S2). These observations are consistent with those from previous analyses for isoform ratio QTLs^22^. Next, we compared the identified sGenes to those assessed in whole blood by the GTEx Consortium^23^ (n=670 individuals), the largest publicly available blood sQTL dataset. Of the 3,013 sGenes discovered by GTEx, 89.0% of the 2,677 we also tested were found as sGenes in our analysis, in addition to 4,470 new sGenes (Table S7). These results demonstrate the value of quantifying *de novo* splicing excision events and the substantially larger sample size.

For a given gene, to test whether corresponding *cis*-eQTLs and *cis*-sQTLs were underpinned by the same genetic variant, we performed colocalization analyses. This revealed 3,979 genes (of 6,252 tested) with colocalized signals (Methods). We found that 49.0% (n=13,490) of tested splicing events had sQTLs that colocalized with an eQTL for the same gene (Table S8). However, of the eQTL-colocalizing splicing events with multiple independent sQTL signals, 82% had additional sQTL loci that did not colocalize with eQTLs. Splicing events with sQTL that did not colocalize with an eQTL were located further downstream of the TSS (median 20.33kb downstream) compared to sQTL signals that did colocalize (median 12.61kb downstream; p=9.8×10^-70^, Wilcoxon test; Figure S3).

### Genetic effects on distal gene expression and splicing

Next, we investigated the distal (*trans*) regulatory effect of genetic variants, defined as >5Mb from the TSS/splicing event. Given the extreme multiple-testing burden for genome-wide *trans*-QTL analyses, we focused on the 53,457 conditionally independent lead *cis*-eSNPs, as these provide a potential mechanism through which a *cis*-acting variant can also affect genes in *trans*.

We identified 2,058 *trans*-eGenes significantly associated with *cis*-eQTLs at the Bonferroni-corrected threshold of p<5×10^-11^ (Figure 2B; Tables S1, S2 and S9). These *trans*-eQTLs were *cis*-eQTLs for 2,498 *cis*-eGenes, and were in *trans* associated with a median number of 3 *trans*-eGenes (range: 1–284). We found that some of them were associated with a large number of *trans*-eGenes, such as *PLAG1* (n=284 genes), *HYMAI* (n=284) and *FUCA2* (n=267). *Cis*-eGenes with a concurrent *trans*-association were significantly enriched for 32 gene ontology (GO) terms, compared to all *cis*-eGenes. Most of the terms related to transcription regulation and immune response, with “metal ion binding” showing the strongest enrichment (p=2.6×10^-30^; Table S10). To further explore these transcriptional regulation mechanisms, we annotated the genes using the Human Transcription Factors database^24^. We found a significant enrichment in sequence-specific transcription factors, representing 14.3% of all *cis*-eGenes with a *trans*-association (357/2,498, p=1.83×10^-38^; Methods). We investigated protein domain annotations for the observed transcription factors and detected a significant enrichment for the C2H2 zinc finger domain (p=9.74×10^-9^ after Bonferroni multiple testing correction), specifically with the Krüppel-associated box (KRAB) domain (p=3.04×10^-10^; Figure S4). For example, the *PLAG1* gene, which is an important regulator of the human hematopoietic stem cell dormancy and self-renewal^25^, codes for a protein with a C2H2 zinc finger domain.

To uncover genetic expression effects impacting distal downstream transcript splicing, we performed a targeted *trans*-analysis using the same 53,457 conditionally independent lead *cis*-eSNPs as in the *trans*-eQTL analysis. The analysis identified significant *trans*-associations for 644 splicing events (209 *trans*-sGenes) at the Bonferroni-corrected threshold of p<8.36×10^-12^. This comprised 758 unique *trans*-sSNPs, corresponding to 566 *cis*-eGenes (Figure 2B; Tables S1 and S11). Of the 644 splicing events regulated in *trans*, 240 (in 91 genes) were not observed to be regulated in *cis*, increasing the total number of splicing events with QTLs. We observed 11 *cis*-eGenes that were implicated by their *cis*-eQTLs in the regulation of more than 10 sGenes in *trans*. For example, we observed that the *cis*-eQTL for the RNA-binding splice factor *QKI* associated with 18 sGenes in *trans* (the most of any eGene; Figure 2C). Across all tissues in GTEx, there were only 29 *trans*-sQTL associations, of which only two were present in whole blood, i.e., the *trans*-splicing of *FYB1* via the *QKI cis*-eQTL, and *trans*-splicing of *ABHD3* for which they did not detect an associated *cis*-effect for the *trans*-sSNP.^23^ Here, we replicated both of these previous *trans*-sGene observations. For *ABHD3*, we demonstrate in addition that this *trans*-sSNP is also a *cis*-eSNP for the splicing factor *TFIP11* and its antisense lncRNA *TFIP11-DT*, potentially regulating the splicing of this gene in *trans*. *Cis*-eGenes of *trans*-sSNPs were significantly enriched for 10 GO terms, including “nucleosome assembly” (p=2.78×10^-6^) and “RNA polymerase II activity (p=1.40×10^-5^) (Table S10).

### Assessment of shared genetic etiology across molecular traits

We next compared transcriptional QTLs to the other -omic trait QTLs derived from subsets of participants from the INTERVAL study. These data include plasma protein QTLs from the Olink Target (“Olink-pQTLs”) and SomaScan panels (“SomaScan-pQTLs”), as well as metabolite QTLs from the Metabolon (“Metabolon-mQTLs”) and Nightingale Health (“Nightingale-mQTLs”) platforms.

To determine whether genetic signals at a given locus across -omic layers reflecting shared genetic or distinct causal variants, we performed statistical colocalization analyses (Methods). This analysis revealed colocalization between either a *cis*-eQTL or -sQTL and *cis*-QTL for 120 Olink-measured proteins (65.9% of analyzed proteins), 404 SomaScan-measured proteins (63.7%), 224 Nightingale-measured metabolites (99.1%) and 495 Metabolon-measured metabolites (81.5%) (Figure 3A; Tables S12–S15). We found colocalized signals across all assessed post-transcriptional molecular phenotypes for 1,229 *cis*-eGenes and 649 sGenes (1,516 unique genes). For Olink- and SomaScan-measured proteins, genetic effect directions were more consistent (p=5.4×10-^10^, Fisher’s exact test) for colocalizing eQTL–pQTL pairs (78.9% with consistent effect directions) than non-colocalizing pairs (59.0%). The uncoupling of eQTLs and pQTLs has previously been observed^26^ and could be due, for example, to post-transcriptional or post-translational mechanisms. Next, we created a network to explore and visualize the interconnectedness among colocalized transcriptional and molecular phenotypes (Figure 3B), linking each phenotype by their colocalizations. For example, we found 7 splicing events in the *OAS1* gene with *cis*-sQTLs that colocalized with both the *cis*-eQTLs for this gene and the OAS1 pQTLs.

**Figure 3.**
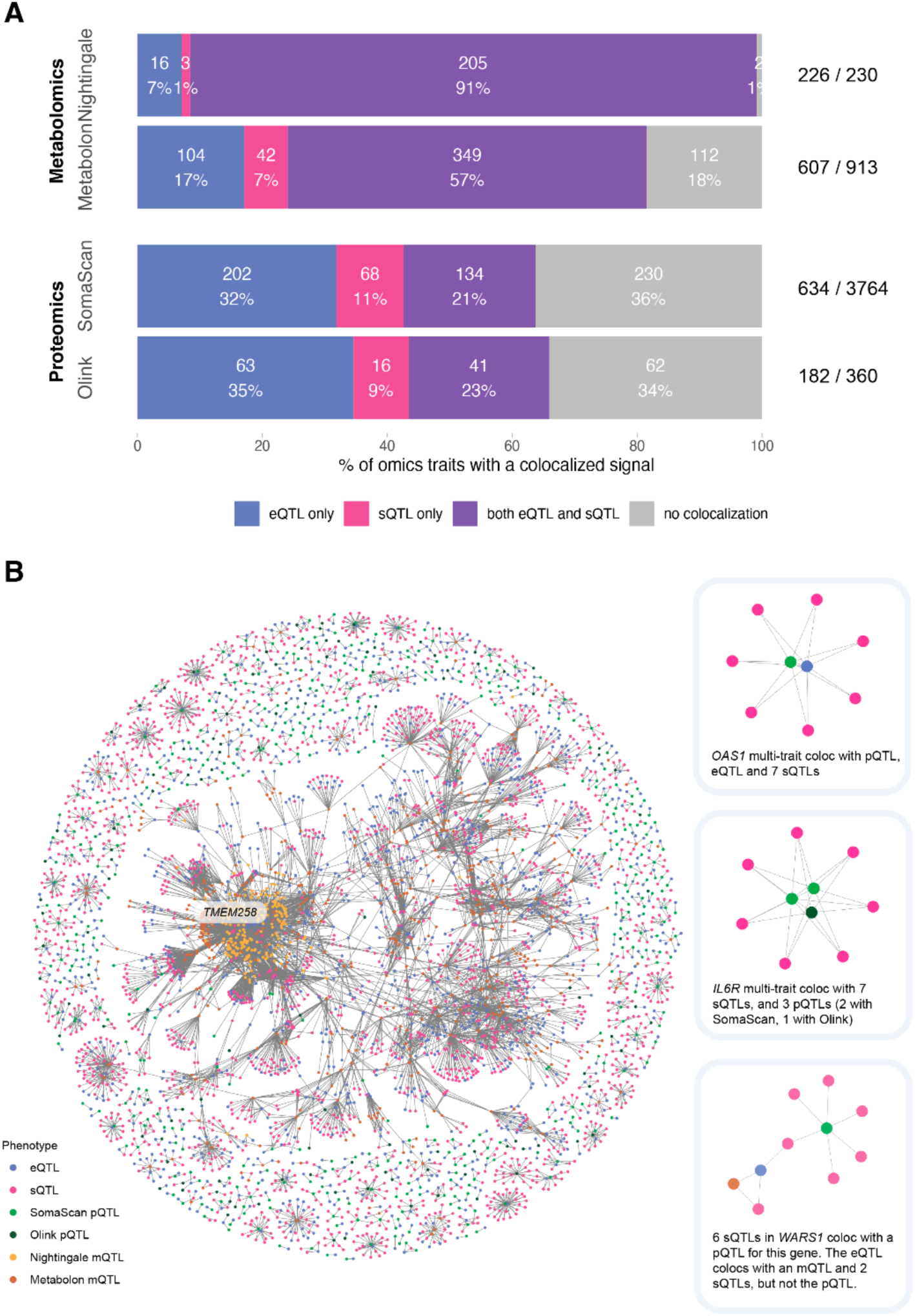
Colocalization analyses of *cis*-eQTL and *cis*-sQTL with other molecular phenotypes. **A)** Barplot of the percentage of -omics traits with a colocalized association signal with a *cis*-eQTL or/and a *cis*-sQTL. **B)** Network graph of all pairwise colocalization results. Highlighted examples on the right-hand side include *OAS1, IL6R,* and *WARS1*.

To investigate the potential mechanisms by which genetic variants impact protein levels through splicing, we annotated the protein domains affected by splicing events. We observed that nearly half of splicing events that colocalized with pQTLs (41.0%, 401 out of 977) excised annotated protein-coding sequences. Splicing has been shown to modulate circulating protein levels through changes in secretion by the inclusion or exclusion of transmembrane domains.^27^ This is exemplified by a splicing event that removes exon 6 of the *FAS* gene, a cell surface receptor for the FAS-ligand (FASL) cytokine. The resulting protein lacking a transmembrane domain is secreted^28^ and competitively inhibits FASL binding, leading to decreased apoptosis. We identified both *cis*-eQTLs for *FAS* and *cis*-sQTLs for this splicing event but these signals were distinct and did not colocalize (max. posterior probability=0.02), and *cis*-sQTLs for excision of the transmembrane domain strongly colocalized with the pQTL (posterior probability=1.00). Similarly, the interleukin-6 and interleukin-7 receptors (IL6R and IL7R, respectively) have previously been reported to produce secreted isoforms through the excision of transmembrane domains.^29,30^ Here, we show that the pQTLs for IL6R and IL7R colocalized with *cis*-sQTLs excising these transmembrane domain-encoding exons, in the absence of *cis*-eQTL colocalization (Figure 3B). This observation emphasizes the role of transcript splicing as a mechanism independent of total transcript abundance through which genetic variation can modify downstream molecular phenotypes. Further, we observed a pQTL colocalizing with an sQTL for the excision of a transmembrane domain in the encoding mRNA in 69 proteins, with 60.2% of these independent sQTL signals (n=100/166) not colocalizing with eQTLs for the same gene (Table S16). For example, this is observed in alpha-1 antitrypsin encoded by *SERPINA1*, and apolipoprotein L1 encoded by *APOL1*.

To maximize statistical power for colocalization, we extended our analyses to the largest available SomaScan- and Olink-pQTL datasets provided by deCODE^8^ (n=35,559 individuals, n=4,719 proteins) and the UK Biobank Pharma Proteomics Project^12^ (UKB-PPP; n=54,306 individuals, n=2,941 proteins), respectively. Focusing on the significant pQTLs overlapping our derived eQTLs and sQTLs, we performed colocalization analysis on 1,608 Olink- and 1,410 SomaScan-measured proteins with our transcriptional phenotypes. This increased the discovery of pQTL-eQTL/sQTL colocalizations from 120 to 1,203 Olink-measured proteins, and from 404 to 984 SomaScan-measured proteins. Additionally, we observed a substantial overlap of eGenes and splicing events with QTLs colocalizing between our internal and the larger external pQTL cohorts. In UKB-PPP, we replicated 95.1% and 79.3% of eQTLs and sQTLs colocalizations respectively, and in deCODE, 87.0% and 80.3% of eQTLs and sQTLs, respectively. These results are summarized in Tables S12-14, and are available to explore in our online resource.

### Mapping causal transcriptional events on downstream molecular phenotypes through mediation analysis

To assess causality of the transcriptional phenotypes on downstream molecular phenotypes, we performed mediation analyses focusing on colocalizing molecular traits assayed in the INTERVAL study (Figure 4A; Methods). The expression of 143 *cis*-eGenes significantly mediated the effect of 413 *cis*-eSNPs on 202 downstream molecular phenotypes, including 101 SomaScan-measured proteins, 54 Olink-measured proteins, 39 Nightingale-measured metabolites, and 8 Metabolon-measured metabolites. In total, this comprised 525 significant eQTL mediation models (variant-gene-molecular phenotype triplets) (Figure 4B). Similarly, we observed 106 splicing event phenotypes in 47 sGenes that significantly mediated the effect of 152 *cis*-sSNPs on 50 downstream molecular phenotypes, including 32 SomaScan-measured proteins, 16 Olink-measured proteins, 1 Nightingale-measured and 1 Metabolon-measured metabolite. In total, this comprised 241 significant sQTL mediation models (Tables S17 and S18; Methods).

**Figure 4.**
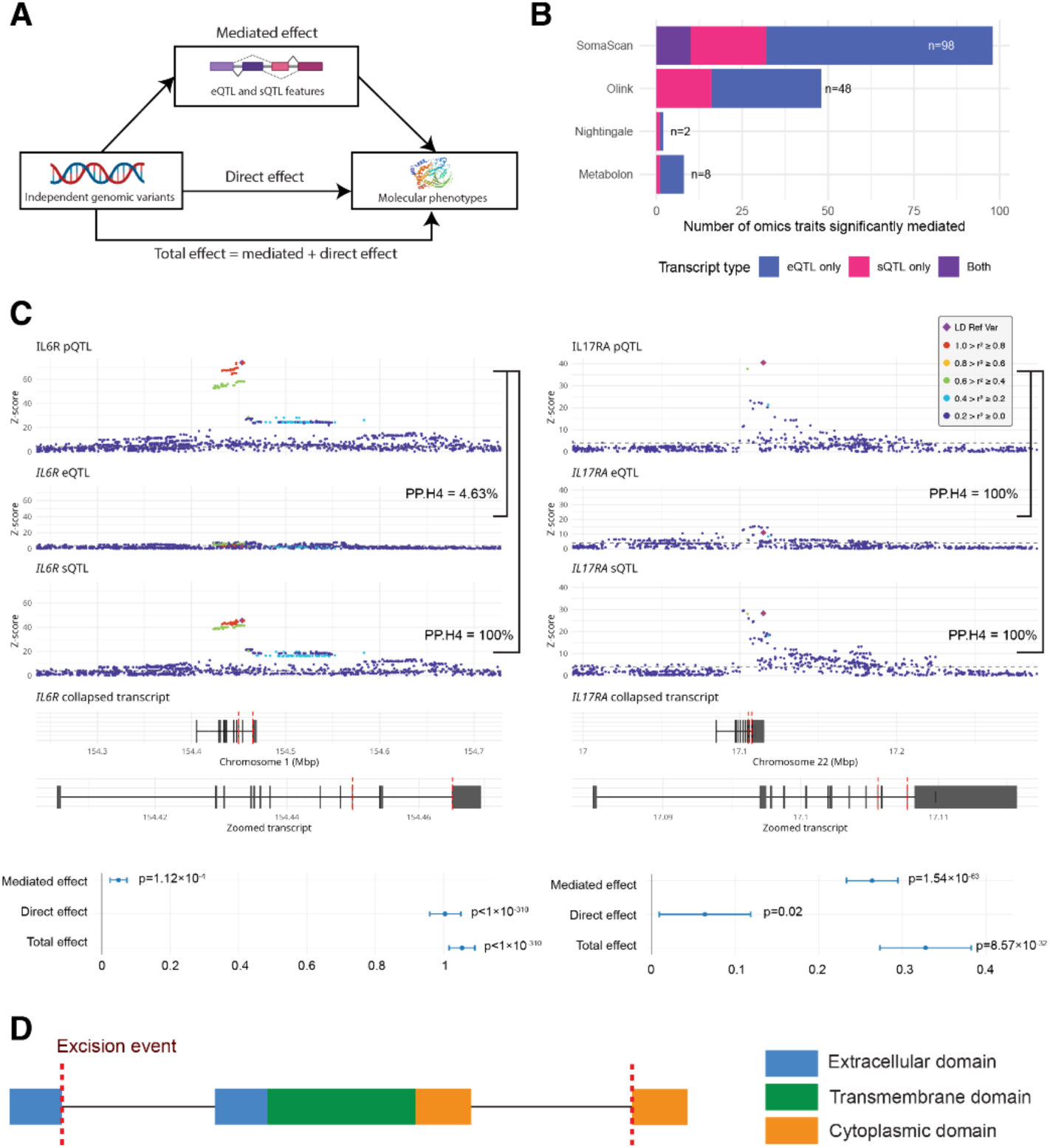
Mediation analyses of molecular phenotypes with transcriptional QTLs. **A)** Schematic of the tested mediation model, for which eQTL and sQTL phenotypes mediate the relationship between genomic variants and levels of molecular phenotypes. **B)** Total number of detected molecular phenotypes mediated by sQTLs and eQTLs. **C)** Colocalization of sQTLs excising the transmembrane domains of the interleukin receptors *IL6R* and *IL17RA*, and mediation with plasma protein quantities. **D)** Schematic of the splicing events excising transmembrane domains of the interleukin receptors *IL6R* and *IL17RA*.

Previous reports have defined the effect of the missense SNP rs2228145 on IL6R ectodomain shedding by ADAM10/17 metalloproteinases, as the variant alters one of their cleavage sites.^31,32^ In line with this finding, we observed the previously mentioned *IL6R* transmembrane splicing event mediating a minority of the effect of a SNP tagging this missense variant (rs12126142; r^2^>0.99; D’>0.99) on Olink-measured plasma protein abundance (4.67%, p=1.12×10^-4^) (Figures 4C and 4D). This suggests a potential dual action of the sSNP or tagged variants on removing this domain and, hence, creating a soluble isoform by both splicing and proteolytic pathways. Conversely, the colocalized signal (lead *cis*-sSNP rs34495746) between splicing of the transmembrane domain of *IL17RA* and levels of its plasma protein was found to have a majority of the effect mediated by transcript splicing (90.41%, p=1.14×10^-43^; Figure 4C). Consistent with this observation, neither the lead SNP nor any strong tagging SNPs (r^2^>0.8) were missense variants.

### Deconvoluting mechanisms of GWAS disease loci with transcriptional and molecular phenotypes

Molecular QTLs can provide insights into the mechanisms underlying genetic variants that influence disease risk.^33^ We performed colocalization analyses with genetic association signals for 20 disease phenotypes from the FinnGen project (release 9)^34^. We prioritized these 20 phenotypes based on their relevance to the circulatory system and available sample size (i.e., ≥1,000 cases; Table S19).

We observed colocalization of disease associated signals with 649 *cis*-eGenes and 365 *cis*-sGenes (1,035 splicing events) across all tested traits (Tables S20 and S21). Many of these independent signals (136/981 for *cis*-eQTLs, 304/1589 for *cis*-sQTLs) also colocalized with the previously tested pQTLs and mQTLs, revealing the regulatory pathways underlying the complex trait-associated variants. For example, a *cis*-sQTL for transmembrane domain splicing of the interleukin-7 receptor (*IL7R*) colocalized with an association locus for dermatitis and eczema, as well as a pQTL for IL7R in UKB-PPP (Figure 5A). This analysis implicates soluble isoforms of IL7R generated by alternative splicing in this condition. The alternative allele of rs6897932 (T) is associated with decreased excision of the *IL7R* transmembrane domain, lower levels of IL7R in plasma and reduced risk of dermatitis and eczema. This allele has been previously shown to associate with decreased lymphocyte count^35^ and decreased risk of multiple sclerosis^36^, suggesting consistent therapeutic implications.

**Figure 5.**
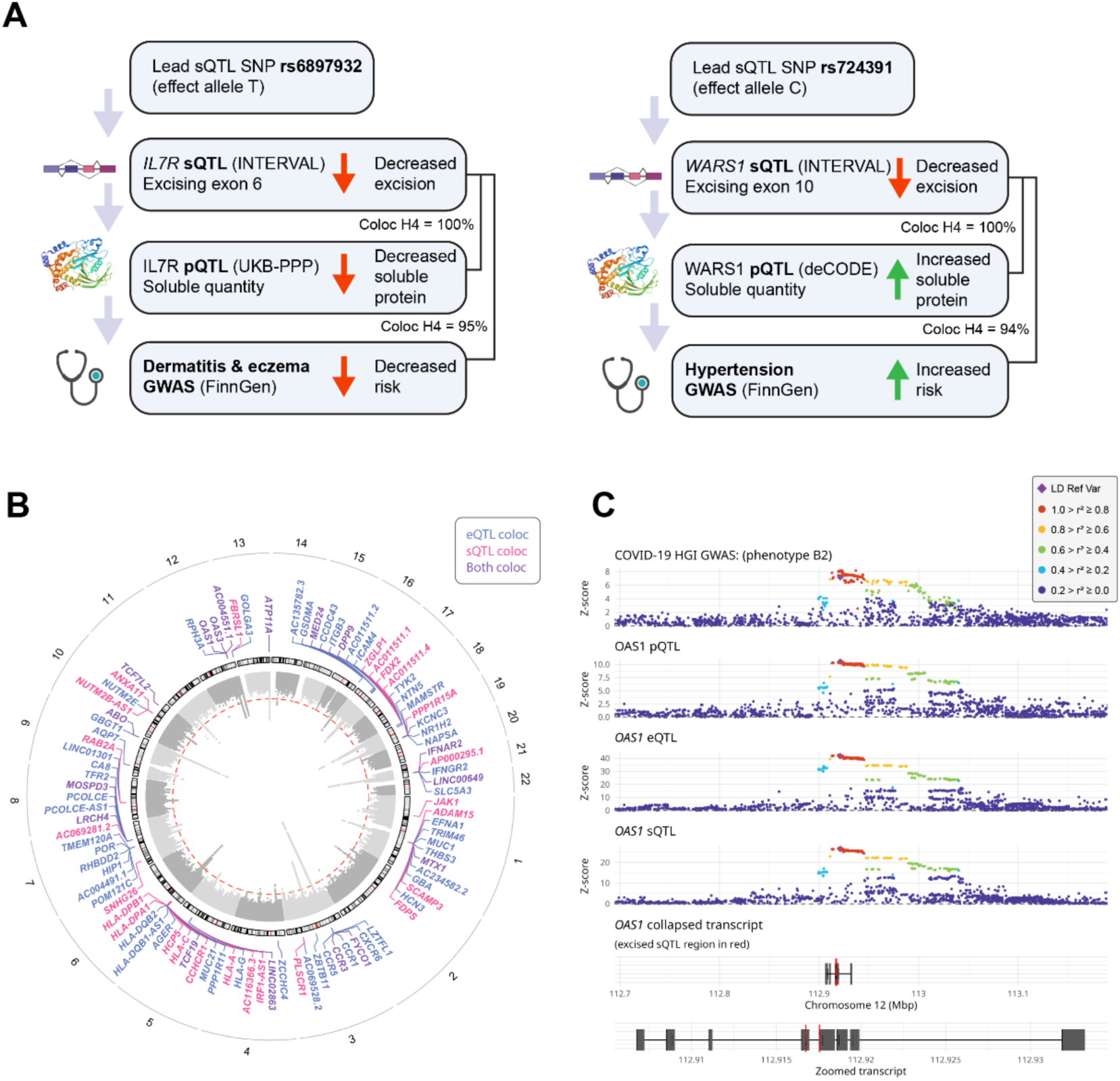
Multi-trait colocalization of *cis*-eQTLs and *cis*-sQTLs with molecular phenotypes and health outcomes. **A)** Putative pathways and directions of effect of sQTL signals for *IL7R* and *WARS1* associated with plasma protein quantity, dermatitis and eczema, and hypertension, respectively. **B)** Gene-level summary of colocalization of *cis*-eQTL and *cis*-sQTL with COVID-19 HGI summary statistics. **C)** Example of a multi-trait colocalization for COVID-19 in *OAS1,* with GWAS summary statistics, *cis*-pQTL, -eQTL, and -sQTL.

Tryptophanyl-tRNA synthetase 1 (encoded by *WARS1*) exists in both secreted and intracellular forms^37^, with downstream impacts on vascular permeability^38^. Here, we found a *cis*-sQTL for excision of exon 10 of *WARS1* (encoding a portion of the tRNA synthetase protein domain), which colocalized with both the WARS1 pQTLs and risk for hypertension in FinnGen (Figure 5A). The alternative allele of rs724391 (C) is associated with decreased excision of exon 10, higher plasma protein levels of WARS1, and increased risk of hypertension.

### Transcriptional mechanisms underlying COVID-19 susceptibility and severity

The large majority of whole-blood RNA is derived from circulating immune cells. Given the importance of the host immune response in COVID-19, we conducted colocalization analyses of the identified eQTLs and sQTLs with genetic loci associated with COVID-19 susceptibility and severity available from the pan-biobank COVID-19 Host Genetics Initiative^39^. We found colocalized signals with COVID-19 loci for 67 *cis*-eGenes and 42 *cis*-sGenes (91 splicing events; Tables S22 and S23; Figure 5B), of which 17 overlapped.

Previous analyses have identified genetic variants that impact splicing of *OAS1*.^40,41^ These variants have subsequently been implicated in influencing COVID-19 severity.^41^ Consistent with these data, we observed colocalization of an eQTL and sQTLs for 7 splicing events at the *OAS1* locus with COVID-19 (Figure 5C). Adjusting for *OAS1* gene expression levels did not ablate the sQTL signals (p<1×10^-16^), suggesting the presence of multiple independent transcriptional mechanisms at this locus. In addition, we found colocalization for these eQTLs and sQTLs with the OAS1 pQTL, suggesting that genetic variants mediate disease risk through transcriptional changes impacting soluble protein levels.

Further, the GWAS signals for COVID-19 susceptibility and severity at the *IFNAR2* locus (encoding the interferon alpha/beta receptor 2) colocalized with a *cis*-eQTL, and *cis*-sQTLs associated with 10 splicing events in this gene. This included a splicing event excising exons 8 and 9, encoding the IFNAR2 transmembrane domain. Rare mutations in exon 9 of this gene leading to loss of function (stop-gain) have been previously reported to increase the risk of severe COVID-19 infection.^42^ While IFNAR2 was not measured by the proteomic assays, isoforms of *IFNAR2* lacking the transmembrane domain are known to generate a soluble protein isoform,^43^ and significantly higher quantities of soluble IFNAR2 have been observed in the serum of patients with severe COVID-19.^44^ However, the role of splicing in this gene on disease severity has not been previously reported. Notably, the colocalizing *IFNAR2* eQTLs are also *trans*-sQTLs for five splicing events in *IFI27*, four of which do not have an association in *cis*. Our results provide evidence for a mechanism whereby common variants regulating splicing of *IFNAR2* could be contributing to disease severity through impacts on protein solubility.

## Discussion

Non-protein-coding genetic variants play an important role in the genetics of complex traits, with 90% of common-trait heritability attributed to this class of variants.^45^ To define the biological mechanisms through which these variants act, additional molecular data and integrative analyses are required. Genome-wide, multi-layered molecular QTL data can help elucidate the functional impact of trait-associated variants and their regulatory networks that underpin complex disease biology. To this end, we discovered eQTLs for 17,233 genes and sQTLs for 29,514 splicing phenotypes in 6,853 genes in peripheral blood through RNA-sequencing of 4,732 individuals. Then, we combined these data with mQTL and pQTL data in the same participants of the INTERVAL study to map the genetic basis for disease phenotypes. All generated data are accessible to the scientific community via the INTERVAL QTL portal (https://IntervalRNA.org.uk) and the cognate OmicsPred portal^46^ (https://www.omicspred.org/).

In comparison to eQTLs, the genetic determinants of splicing have been less thoroughly explored; in particular, how they impact on downstream molecular phenotypes and disease risk. Our data support previous findings that splicing QTLs are major contributors to complex traits.^47^ Through mapping sQTLs alongside eQTLs, we identified additional independent mechanisms by which genetic variants can influence mRNA and protein levels. For example, 98 splicing events that colocalized with pQTLs (such as IL6R and IL7R) excised protein-coding sequences encoding transmembrane domains. Many of these pQTLs did not colocalize with eQTLs, suggesting that the sQTLs provide the pivotal mechanistic insight, given that genetic effects on splicing are more highly shared between tissues than genetic effects on expression^23^. Furthermore, by identifying and utilizing *de novo* excision events from the RNA-seq data, we increased the resolution beyond established transcript annotations.

Using the multi-omic data in the INTERVAL study, we systematically performed mediation analyses to evaluate causality in the context of colocalized genetic association signals with molecular traits. In total, we observed 222 molecular phenotypes significantly mediated by gene expression or splicing, providing an additional layer of evidence to delineate functional mechanisms. For instance, we found that an sQTL excising the extracellular domain of *CD33* mediated the majority of the effect of the sSNP on CD33 soluble protein levels. Mediation analyses are important to define the mode-of-action of the genetic effects underlying association loci identified in GWAS, as well as the magnitude and direction of their relative effects on downstream phenotypes.

Our study has limitations. First, statistical power was limited to map genome-wide eQTLs and sQTLs in *trans*. As *trans*-QTLs are challenging to replicate and distinguish from cell type heterogeneity in bulk RNA-seq studies,^9^ we prioritized the identified conditionally independent lead eSNPs for our *trans*-QTL analyses to prioritize the mechanism of upregulated gene expression modifying the expression and splicing of downstream genes. Large-scale meta-analyses of *trans*-QTL datasets will be required to create a resource of replicated associations, such as that being prepared by the eQTLGen Consortium (https://www.eqtlgen.org/). Second, our analyses comprised proteins quantified in plasma, rather than intracellular proteins. Thus, the interpretation of the effects of gene expression and splicing QTLs on proteins may be due to impacts on both quantity and solubility, and other regulatory mechanisms may not be captured. Third, the intrinsic properties of the different molecular data types can create challenges in interpretation. For example, there is considerable correlation structure between metabolite levels.^48^ As such, we found that the majority of mQTLs (96%) colocalized with either a *cis*-eQTL or *cis*-sQTL. Conversely, mQTLs showed mediation by *cis*-eQTL or *cis*-sQTLs less frequently than pQTLs (i.e., 6.8% vs 32.6% for mQTLs and pQTLs, respectively). Lastly, our cohort comprised individuals of European ancestry. More work is needed to establish the translatability of our findings to other ancestries.

Previous studies showed that local regulation of gene expression is largely shared across tissues,^49^ and that larger, well-powered eQTL studies in a surrogate tissue may identify more trait-colocalizing eQTLs than smaller studies in the target tissue.^50^ Hence, these results provide a scientific rationale for the generation of increasingly large-scale QTL data in easily accessible tissues, such as peripheral blood. In our study, we further demonstrate the value of such a dataset when integrating data from multiple molecular phenotypes in the same individuals and linking these to external health outcomes to help address the variant-to-function challenge (e.g., through statistical colocalization and mediation analyses). Similar application to population biobanks is warranted and with the emerging availability of concomitant molecular data at the single-cell level across a wide range of tissues, single-cell-QTL mapping at population scale will become feasible. Such data will enable us to dissect gene regulatory networks at much greater resolution across specific cell types and dynamic processes.^51,52^ Our genetic discoveries are publicly available in an open-access and interactive resource at https://IntervalRNA.org.uk.

## Acknowledgements

Participants in the INTERVAL randomized controlled trial were recruited with the active collaboration of NHS Blood and Transplant England (https://www.nhsbt.nhs.uk/), which has supported field work and other elements of the trial. DNA extraction and genotyping were co-funded by the National Institute for Health and Care Research (NIHR), the NIHR BioResource (https://bioresource.nihr.ac.uk/) and the NIHR Cambridge Biomedical Research Centre (BRC-1215-20014) [*]. RNA-seq was funded as part of an alliance between the University of Cambridge and the AstraZeneca Centre for Genomics Research, and by the NIHR Cambridge Biomedical Research Centre (BRC-1215-20014) [*]. Olink Target assays (Neurology panel) were funded by Biogen, Inc. SomaLogic assays were funded by Merck & Co, Inc and the NIHR Cambridge Biomedical Research Centre (BRC-1215-20014) [*]. Metabolon HD4 assays were funded by the NIHR BioResource; NIHR Cambridge Biomedical Research Centre (BRC-1215-20014) [*]; Wellcome Trust grant number 206194; and BioMarin Pharmaceutical, Inc. Nightingale Health assays were funded by the European Commission Framework Programme 7 (HEALTH-F2-2012-279233). The academic coordinating center for INTERVAL was supported by core funding from the NIHR Blood and Transplant Research Unit (BTRU) in Donor Health and Genomics (NIHR BTRU-2014-10024); NIHR BTRU in Donor Health and Behaviour (NIHR203337); UK Medical Research Council (MR/L003120/1); British Heart Foundation (SP/09/002; RG/13/13/30194; RG/18/13/33946); and NIHR Cambridge BRC (BRC-1215-20014; NIHR203312) [*]. A complete list of the investigators and contributors to the INTERVAL trial is provided in Di Angelantonio *et al*.^16^ The academic coordinating center would like to thank blood donor center staff and blood donors for participating in the INTERVAL trial. This work was supported by Health Data Research UK, which is funded by the UK Medical Research Council, Engineering and Physical Sciences Research Council, Economic and Social Research Council, Department of Health and Social Care (England), Chief Scientist Office of the Scottish Government Health and Social Care Directorates, Health and Social Care Research and Development Division (Welsh Government), Public Health Agency (Northern Ireland), British Heart Foundation and Wellcome. *The views expressed are those of the authors and not necessarily those of the NIHR or the Department of Health and Social Care. The Wellcome Sanger Institute is supported by core funding from the Wellcome Trust (206194 and 220540/Z/20/A). We thank the Wellcome Sanger Institute’s Scientific Operations team for their contribution to sequencing data generation. For the purpose of Open Access, the authors have applied a CC BY public copyright licence to any Author Accepted Manuscript version arising from this submission. This work was performed using resources provided by the Cambridge Service for Data Driven Discovery (CSD3) operated by the University of Cambridge Research Computing Service (https://www.csd3.cam.ac.uk/), provided by Dell EMC and Intel using Tier-2 funding from the Engineering and Physical Sciences Research Council (capital grant EP/P020259/1), and DiRAC funding from the Science and Technology Facilities Council (https://dirac.ac.uk/). We thank the participants and investigators of the UK Biobank study who made this work possible (Resource Application Numbers 26041 and 65851).

## Personal funding/acknowledgements

- A.T. is supported by the Wellcome Trust (PhD studentship 222548/Z/21/Z).
- E.P. was funded by the EU/EFPIA Innovative Medicines Initiative Joint Undertaking BigData@Heart grant 116074 and is funded by the NIHR BTRU in Donor Health and Behaviour (NIHR203337) [*].
- M.A.Q. is on the KOL panel for New England Biolabs.
- S.C.R was funded by a BHF Programme Grant (RG/18/13/33946) and the NIHR Cambridge BRC (BRC-1215-20014; NIHR203312).
- B.B.S. and H.R. are employees and stockholders of Biogen.
- Y.X. is supported by the UK Economic and Social Research Council (ES/T013192/1).
- C.D.W. is an employee and stockholder of Johnson & Johnson.
- S.P. and D.S.P. are employees and stockholders of AstraZeneca.
- D.J.G. is an employee and stockholder of BioMarin Pharmaceutical.
- D.J.R. is an employee of NHS Blood and Transplant.
- J.E.P. is supported by a Medical Research Foundation Fellowship (MRF-057-0003-RG-PETE-C0799) and has received hospitality and travel expenses to speak at Olink-sponsored academic meetings (none within the past 5 years).
- J.D. holds a British Heart Foundation Professorship and a NIHR Senior Investigator Award [*].
- A.S.B. has received grants outside of this work from AstraZeneca, Bayer, Biogen, BioMarin and Sanofi.
- M.I. is supported by the Munz Chair of Cardiovascular Prediction and Prevention and the NIHR Cambridge Biomedical Research Centre (BRC-1215-20014; NIHR203312) [*]. M.I. is also supported by the UK Economic and Social Research Council (ES/T013192/1). M.I. is a trustee of the Public Health Genomics (PHG) Foundation, a member of the Scientific Advisory Board of Open Targets, and has a research collaboration with AstraZeneca that is unrelated to this study.

## Materials and Methods

### Study participants

The INTERVAL study is a prospective cohort study of approximately 50,000 participants nested within a randomized trial of varying blood donation intervals.^15,16^ Between 2012 and 2014, blood donors aged 18 years and older were recruited at 25 centers of England’s National Health Service Blood and Transplant (NHSBT). All participants gave informed consent before joining the study and the National Research Ethics Service approved this study (11/EE/0538). Participants were generally in good health as blood donation criteria exclude individuals with a history of major diseases (e.g. myocardial infarction, stroke, cancer, HIV, and hepatitis B or C) and who have had a recent illness or infection. Participants completed an online questionnaire comprising questions on demographic characteristics (e.g. age, sex, ethnicity), lifestyle (e.g. alcohol and tobacco consumption), self-reported height and weight, diet and use of medications.

### Blood collection

Blood samples were collected from all INTERVAL participants at baseline and also from ∼60% of participants approximately 24 months after baseline. For a subset of ∼5,000 participants at the 24-month time point, an aliquot of 3 ml of whole blood was collected in Tempus Blood RNA Tubes (ThermoFisher Scientific), following the manufacturer’s instructions, and then transferred at ambient temperature to the UK Biocentre (Stockport, UK). Samples were stored at -80°C until use.

### RNA extraction

RNA extraction was performed by QIAGEN Genomic Services using QIAGEN’s proprietary silica technology. The quality control of the extracted RNA was performed by spectrophotometric measurement on an Infinite 200 Microplate Reader (Tecan). RNA Integrity Number (RIN) values were determined using a TapeStation 4200 system (Agilent), following the manufacturer’s protocol. Samples with a concentration <20 ng/μl and a RIN value <4 were excluded from further analyses.

### Automated RNA-seq library preparation

Samples were quantified with a QuantiFluor RNA System (Promega) using a Mosquito LV liquid handling platform (SPT Labtech), Bravo automation system (Agilent) and FLUOstar Omega plate reader (BMG Labtech), and then cherry-picked to 200 ng in 50 μl (= 4 ng/μl) using a liquid handling platform (Tecan Freedom EVO). Next, mRNA was isolated using a NEBNext Poly(A) mRNA Magnetic Isolation Module (NEB) and then re-suspended in nuclease-free water. Globin depletion was performed using a KAPA RiboErase Globin Kit (Roche). RNA library preparation was done using a NEBNext Ultra II DNA Library Prep Kit for Illumina (NEB) on a Bravo NGS workstation automation system (Agilent). PCR was performed using a KapaHiFi HotStart ReadyMix (Roche) and unique dual-indexed tag barcodes on a Bravo NGS workstation automation system (Agilent). We applied the following PCR programme: 45 sec at 98°C, 14 cycles of 15 sec at 98°C, 30 sec at 65°C and 30 sec at 72°C, followed by 60 sec at 72°C. Using a Zephyr liquid handling platform (PerkinElmer), PCR products were purified using AMPure XP SPRI beads (Agencourt) at a 0.8:1 bead:sample ratio and then eluted in 20 μl of Elution Buffer (QIAGEN). RNA-seq libraries were quantified with an AccuClear Ultra High Sensitivity dsDNA Quantitation Kit (Biotium) using a Mosquito LV liquid handling platform (SPT Labtech), Bravo automation system (Agilent) and FLUOstar Omega plate reader (BMG Labtech). Then, libraries were pooled up to 95-plex in equimolar amounts on a Biomek NX-8 liquid handling platform (Beckman Coulter), quantified using a High Sensitivity DNA Kit on a 2100 Bioanalyzer (Agilent), and then normalized to 2.8 nM prior to sequencing.

### RNA sequencing and data pre-processing

Samples were sequenced using 75 bp paired-end sequencing reads (reverse stranded) on a NovaSeq 6000 system (S4 flow cell, Xp workflow; Illumina). The sequencing data were de-plexed into separate CRAM files for each library in a lane. Adapters that had been hard-clipped prior to alignment were reinserted as soft-clipped post alignment, and duplicated fragments were marked in the CRAM files. The data pre-processing, including sequence QC, and STAR and alignments was performed with the Nextflow pipeline publicly available at https://github.com/wtsi-hgi/nextflow-pipelines/blob/rna_seq_interval_5591/pipelines/rna_seq.nf, including the specific aligner parameters. We assessed the sequence data quality using FastQC v0.11.8. Samples mismatched between RNA-seq and genotyping data within the cohort were identified using QTLtools MBV v1.2^53^. Reads were aligned to the GRCh38 human reference genome (Ensembl GTF annotation v99) using STAR v2.7.3a^54^. The STAR index was built against GRCh38 Ensembl GTF v99 using the option - sjdbOverhang 75. STAR was run in a two-pass setup with standard ENCODE options to increase mapping accuracy: (i) a first alignment step of all samples was used to discover novel splice junctions; (ii) splice junctions of all samples from the first step were collected and merged into a single list; (iii) a second step realigned all samples using the merged splice junctions list as input. We used featureCounts v2.0.0^55^ to obtain a count matrix.

### Gene expression quantification

The raw gene-level count data contained N=60,676 genes across N=4,778 individuals with 2.03–95.55 million uniquely mapped reads (median: ∼24 million). Sequencing was performed across 15 batches.

### Quality control of gene expression data

We filtered samples of poor quality by removing samples with a read depth below 10 million uniquely mapped reads. A relatedness matrix was obtained using the PLINK v1.9^56^ -make-rel ‘square’ command on pruned genotype data, and a cut-off threshold of 0.1 was used to define related individuals. For each pair of related individuals, one individual was arbitrarily removed. After quality control, a total of N=46 samples were removed. After the sample QC, we filtered lowly expressed genes by retaining genes with >0.5 counts per million (CPM) in ≥1% of the samples, in line with the filter applied by the eQTLGen consortium^9^. In our dataset, a CPM value of 0.5 roughly equates to having 5 counts in a sample with the lowest read depth (10 million uniquely mapped reads) or 47 counts in a sample with the highest read depth (94 million reads). We further excluded globin genes, rRNA genes, and pseudogenes. After quality control, the final gene expression dataset included 19,173 autosomal genes (13,874 of which are protein-coding) across a total of 4,732 individuals.

### Normalization of gene expression data

Prior to the eQTL analysis, the count data was normalized using trimmed mean of M-values (TMM)^57^ implemented in the R package edgeR v3.24.3. The TMM-normalized values were further converted into fragments per kilobase of transcript per million mapped reads (FPKM) values (log_2_-transformed) to take gene length into account. Next, for each gene, the normalized log_2_-FPKM values across samples were transformed via the ranked-based inverse normal transformation function “rntransform” implemented in the R package GenABEL v1.8-0^58^. Inverse normal transformation was applied to ensure the expression values followed a normal distribution.

### Splicing data generation

Splice junctions were extracted from aligned RNA-seq bams for the 4,732 individuals using regtools v0.5.2^59^ junctions extract (parameters: “-s 1 -m 50”). Introns represented by extracted splice junctions were then clustered into groups based on overlapping start or end sites, with the Leafcutter pipeline v0.2.9^60^ (leafcutter_cluster_regtools.py, parameters: “-m 100 -M 50 -l 100000 - p 0.01”). Clustered introns were then prepared for sQTL analysis with Leafcutter prepare_phenotype_table.py to convert intron counts to normalized ratios and compute 10 splicing PCs. Introns were matched to regions of Ensembl v99 genes and protein domains annotated using a custom pipeline (described in Data Availability). Total observed introns (n=956,722) were filtered to those that were autosomal, overlapping an expressed gene body, with CPM>0.5 in at least 24 individuals, and sufficient variance (minimum 2 filtered splice event phenotypes per cluster), resulting in 111,937 filtered splicing event phenotypes, in 11,016 genes (see Figure S5 for a summary of splicing event QC).

### DNA extraction, genotyping and imputation

In brief, DNA extracted from buffy coat samples collected from INTERVAL participants at the study baseline was used to assay approximately 830,000 variants on the Affymetrix Axiom UK Biobank genotyping array.^61^ Genotyping and sample QC were performed as previously described.^61^ Prior to imputation, additional variant filtering steps were performed to establish a high-quality imputation scaffold including 654,966 autosomal, non-monomorphic, bi-allelic variants with Hardy-Weinberg Equilibrium (HWE) p>5×10^-6^, with a call rate of >99% across the INTERVAL genotyping batches in which a variant passed QC, and a global call rate of >75% across all INTERVAL genotyping batches. Next, variants were phased using SHAPEIT3 and imputed using a combined 1000 Genomes Phase 3-UK10K reference panel. Imputation was performed via the Sanger Imputation Server (https://imputation.sanger.ac.uk) and resulted in 87,696,888 imputed variants. For the present analysis, imputed genotypes were lifted over to reference build GRCh38 using CrossMap v0.3.4^62^ and the Ensembl chain file provided with the package. Imputed genotypes were hard-called with PLINK v2.00a2-32-bit^56^ using the default parameters. Prior to analysis, the dataset was restricted to individuals with RNA-seq, and filtered to remove genetic variants with HWE exact test p<1×10^-6^, genotype missingness >0.05, or MAF <0.5%.

### Identification of sample swaps and cross-contamination

The Match Bam to VCF (MBV) method from QTLTools^53^ was used to identify sample mix-ups and cross-contamination. MBV directly compares each aligned RNA-seq BAM file to all the genotypes in the VCF file and computes the proportion of concordant heterozygous and homozygous sites. To reduce computation time, we only focused on chromosome 1. Based on the concordance (close to 100%) between the genotype data and RNA-seq samples, we identified and corrected for 10 pairs of mislabeled samples. We removed 7 RNA-seq samples that did not show a clear high concordance (highest was <50%) with any particular genotype sample – either due to cross-contamination or the actual matching genotypes were not available. In addition, principal component (PC) analysis was performed to determine whether the 10 pairs of samples were mislabeled in the technical covariate file. We linked PC3 with RIN values to confirm that RIN and other related technical covariates were recorded after the swap occurred. Hence, we corrected the sample IDs for the 10 pairs of mislabeled samples in both the count data and technical covariate file. We linked PC4 with sex information to confirm that sex and other biological covariates we recorded before the swap and did not require correction for mislabeling.

### PEER factor analysis

We used the probabilistic estimation of expression residuals (PEER) method^63^, implemented in the R package peer v.1.0 (downloaded from https://github.com/PMBio/peer), to detect and correct eQTL mapping for latent batch effects and other unknown confounders. PEER factors were estimated while accounting for age, sex, BMI, and 19 blood cell traits (Table S24) as known confounders. PEER was run for 50 factors, converging at 148 iterations. For inclusion in the eQTL analysis, we selected the number of PEER factors based on two criteria: (i) discovery of the largest number of *cis*-eGenes and (ii) additional gain in *cis*-eGenes with incremental increase in PEER factors (Figure S6). We found that the relationship between the increase in number of discovered *cis*-eGenes and incremental increase in PEER factors is similar to that observed in the GTEx whole-blood dataset^23^. Therefore, we included 35 PEER factors in our eQTL analysis, consistent with GTEx.

### Mapping of eQTLs and sQTLs

Expression QTLs and splicing QTLs were called using tensorQTL v1.0.6^64^. The covariates integrated in the regression model are listed and described in Table S24 and S25. In brief, these included (1) demographic variables such as age at blood sampling, sex, and BMI at baseline (since it was not collected at time of blood sampling), (2) technical variables such as RIN, read depth, and season of blood sampling, (3) 10 genotype PCs and 35 PEER factors (for eQTLs) or 10 splicing PCs (for sQTLs) and (4) 19 different blood cell traits. For the *cis*-eQTL analysis, variants were defined as being in *cis* with a gene if they were located within a window of ±1Mb from the TSS. For the sQTL analysis, the window was set to ±500kb from the center of the splicing event to balance primary and secondary sQTL discovery. Feature annotation, including TSS position, was obtained from Ensembl v99 (January 2020). For both *cis*-eQTL and *cis*-sQTL analyses, multiple testing correction was applied in tensorQTL as follows: (1) for each gene (or splicing event), the adjusted lowest p-value was estimated using a beta distribution approximation from a permutation procedure (10,000 permutations)^65^; (2) Benjamini-Hochberg FDR correction was applied to the beta-approximated p-values across genes (or splicing events) and the FDR q-value threshold was set to 5%. For each significant gene (or splicing event), a nominal p-value threshold was estimated to identify significant SNPs. Conditional analysis was performed for each *cis*-eGene (or splicing event phenotype with a *cis*-sQTL) using GCTA-COJO v1.94.0beta (January 2022)^66,67^. The program took as input the gene *cis*-eQTL (or *cis*-sQTL) summary statistics, the INTERVAL imputed genotype data for *cis*-variants and the p-value threshold used to identify the *cis*-eGene (or splicing QTL). A *trans*-eQTL analysis was performed on the list of lead SNPs from *cis*-eGenes independent signals. The *trans*-regions were defined as genomic regions outside of the ±5Mb window from the TSS. The Bonferroni multiple-testing correction method (i.e., p=0.05/number of tested *trans*-associations) was applied to identify significant *trans*-associations.

### Validation of *cis*-eQTL and *cis*-sQTL results

Results from the *cis*-eQTL analysis were compared to the results obtained in the eQTLGen study^9^, which are available at https://www.eqtlgen.org/cis-eqtls.html. In our comparison, we explored the percentage of overlap of *cis*-eGenes and the effect direction of genetic associations. For the overlap of *cis*-eGenes, we focused on the list of 15,722 genes that were tested in both INTERVAL and eQTLGen. For the comparison of effect directions, we computed the correlation of Z-scores for SNPs that were the most significant in INTERVAL for each gene and that were also tested in eQTLGen. Results from sQTL analysis were compared in the same way to sGenes discovered in GTEx whole blood sQTLs (version 8)^23^, which are available at https://gtexportal.org/home/datasets.

### Enrichment analyses

Enrichment analyses were performed using a one-sided Fisher’s exact test, on QTL results annotated with GO terms^68^ (downloaded in May 2022) and the Human Transcription Factors database^24^. We tested for enrichment within *cis*-eGenes with a *trans*-association with gene expression or splicing using significant *cis*-eGenes as background.

### Colocalization analysis

Colocalization analysis was performed using the results of conditional analysis from GCTA-COJO^66,67^ and the R package coloc v5.1.0.1^69^ on pairwise independent QTL signals following the pwCoCo methodology^70^. The colocalization analysis window was the entire *cis*-window, i.e., ±1Mb for eQTLs and ±500kb for sQTLs. Prior probabilities were kept as the default values, i.e., p1=1×10^-4^, p2=1×10^-4^, p12=1×10^-5^. Colocalized results were defined with the thresholds PP3+PP4>=0.9 and PP4/PP3>=3, PP3 and PP4 being the posterior probabilities of hypotheses 3 and 4 as outlined previously^69^. For colocalization analysis with external omics data, summary statistics were downloaded from each study (Table S26 for the description of the different -omics studies). A previous study performed simulations showing that the impact of complete sample overlap on colocalization results was negligible with large sample sizes.^71^ Prior to colocalization analysis, (1) proteins were annotated using the R package biomaRt v2.46.3 to obtain corresponding genes in Ensembl v99 (January 2020); (2) significant pQTLs and mQTL were filtered. For pQTLs, p-value thresholds per feature were defined by a two-step multiple testing correction^72,73^. For mQTLs, we used a Bonferroni-adjusted p-value threshold of p<5×10^-8^, corrected for the number of metabolites analyzed.

### Mediation analysis

Mediation analyses were conducted using the natural effects model implemented in the R package medflex v0.6-7^74^. In the models, we defined (1) the independent lead eQTL (or sQTL) SNP (coded as 0, 1 and 2) as the independent (exposure) variable, (2) the gene expression level (or splicing event phenotype) as the mediator and (3) the molecular trait as the dependent (outcome) variable. Gene expression (or splicing event phenotype) residuals were computed after adjusting for the same covariates as we used for eQTL/sQTL mapping, while molecular traits were adjusted for covariates described by each study (Table S26). For all mediation analyses, samples with missing genotype or molecular data were removed. Standard errors were computed based on the robust sandwich estimator. Significant direct, indirect and total effects were identified after Bonferroni multiple-testing correction between each molecular phenotype assay.

### Interactive QTL web-portal

To facilitate accessibility of the results, a web-portal was built to enable exploration of eQTL and sQTLs. Summary statistics and expression phenotypes were imported into a MariaDB v10.2.38 database, and code written to facilitate their retrieval in PHP v7.2.34 with jquery v3.5.1, and styled with Bootstrap v3.4.1. Tables are powered by DataTables v1.13.3, locus plots are visualized with LocusZoomJS v0.13.4, and QTL plots with plotly v2.9.0.

## Data Availability

The INTERVAL study data used in this paper are available to bona fide researchers from. The data access policy for the data is available at http://www.donorhealth-btru.nihr.ac.uk/project/bioresource. The newly generated RNA-sequencing data (n=4,732 INTERVAL participants) have been deposited at the European Genome-phenome Archive (EGA) under the accession number EGAD00001008015. The results from the genetic association, colocalization and mediation analyses are available at https://IntervalRNA.org.uk. All original code has been deposited at GitHub at https://github.com/INTERVAL-RNAseq/manuscript-scripts.

## Supplementary Tables

**Table S1: Summary of eQTL and sQTL results.**

**Table S2. List of genes for cis- and trans-eQTL analyses.** Each of 19,173 genes was annotated whether it was tested and a significant association was found from the *cis*- and/or *trans*-eQTL mapping analysis. *Cis*-eQTL significance information was also retrieved for the eQTLGen study. Headers are: feature_id: gene ID from Ensembl v99; gene_name: gene name (Ensembl v99); gene_biotype: gene biotype (Ensembl v99); *cis*_eQTL_tested: indicates if the gene has been tested in *cis*-eQTL analysis (1 if tested, 0 otherwise); *cis*_eQTL_significant: indicates if the gene is a significant *cis*-eGene (1 if significant, 0 otherwise); *cis*_eQTL_significant_with_*trans*: indicates if the *cis*-eGene has eSNPs or lead independent SNPs in *trans*-association with another gene; *trans*_eQTL_tested: indicates if the gene has been tested in *trans*-eQTL analysis (1 if tested, 0 otherwise); *trans*_eQTL_significant: indicates if the gene is a significant *trans*-eGene (1 if significant, 0 otherwise); *cis*_eQTL_tested_in_eQTLGen: indicates if the gene has been tested in the *cis*-eQTL mapping analysis from eQTLGen study (1 if tested, 0 otherwise); *cis*_eQTL_significant_in_eQTLGen: indicates if the gene is a significant *cis*-eGene in the eQTLGen study (1 if significant, 0 otherwise).

**Table S3. Independent signals per cis-eGene after conditional analysis.** This table contains information for the total of *cis*-eQTL 56,959 independent association signals. Headers are: phenotype_id: gene ID (Ensembl v99); gene_name: gene name (Ensembl v99); variant_id: variant ID (rsid or chr_pos_A1_A2); chr: chromosome; pos_b38: genomic position of the variant for the GRCh38 assembly; tss_distance: genomic distance between the variant and the TSS in bp for the GRCh38 assembly; variant_id_b37: variant ID for the GRCh37 assembly; pos_b37: genomic position of the variant for the GRCh37 assembly; effect_allele: effect allele; other_allele: other allele; af: allele frequency of the effect allele; slope: effect size of the effect allele; slope_se: standard error for the effect size; pval_nominal: nominal p-value; bJ: effect size from a joint analysis of all the selected SNPs from GCTA-cojo; bJ_se: standard error of the effect size from the joint analysis; pJ: p-value from the joint analysis; LD_r: LD correlation between the SNP i and SNP i + 1 for the SNPs on the list; rank: rank of the independent association signal.

**Table S4. Summary of the comparison of cis-eQTL results with eQTLGen study.**

**Table S5. List of splicing events with a cis-sQTL.** This table contains annotation for each of the 29,514 splicing events with a *cis*-sQTL. Headers are: phenotype_id: splicing ID (chr:start:end:cluster_id_strand); splice_clu: splicing cluster ID; range: genomic range of the excised intron; qval: FDR-adjusted nominal p-value; variant_rsid: lead variant ID; af: allele frequency of the effect allele of the lead variant; z_score: Z-score of the lead variant; gene_name: gene name(s) (Ensembl v99), “(OS)” after the gene name indicates that the splicing event is mapped on the opposite strand of the gene; gene_id: gene ID(s) (Ensembl v99), “(OS)” after the gene ID indicates that the splicing event is mapped on the opposite strand of the gene; gene_biotype: gene biotype from Ensembl v99; os: True if on opposite strand to annotated gene; ov_cds: spliced range overlaps a CDS region (Gencode 33); ov_exon: spliced range overlaps a known exon (Gencode 33); unipLocTransMemb: Spliced range overlaps a transmembrane domain (Uniprot); unipLocCytopl: spliced range overlaps a cytoplasmic domain (Uniprot); unipLocExtra: spliced range overlaps an extracellular domain (Uniprot); unipLocSignal: spliced range overlaps a signal peptide domain (Uniprot); unipDomain: spliced range overlaps any annotated protein domain (Uniprot); pfamDomain: spliced range overlaps any annotated protein domain (Pfam); P5_exon: spliced range matches known 5’ exon boundary (Gencode 33); P3_exon: spliced range matches known 3’ exon boundary (Gencode 33); P5_p3: spliced range matches known 5’ AND 3’ exon boundaries (Gencode 33); P5_or_p3: spliced range matches known 5’ OR 3’ exon boundary (Gencode 33).

**Table S6. Independent signals per cis-sQTL after conditional analysis.** This table contains information for the total of *cis*-sQTL 47,050 independent association signals. Headers are: phenotype_id: splicing event ID (chr:start:end:cluster_id_strand); gene_id: gene ID(s) (Ensembl v99); gene_name: gene name(s) (Ensembl v99); variant_id: variant ID (rsid or chr_pos_A1_A2); chr: chromosome; pos_b38: genomic position of the variant for the GRCh38 assembly; tss_distance: genomic distance between the variant and the TSS in bp for the GRCh38 assembly; variant_id_b37: variant ID for the GRCh37 assembly; pos_b37: genomic position of the variant for the GRCh37 assembly; effect_allele: effect allele; other_allele: other allele; af: allele frequency of the effect allele; slope: effect size of the effect allele; slope_se: standard error for the effect size; pval_nominal: nominal p-value; bJ: effect size from a joint analysis of all the selected SNPs from GCTA-cojo; bJ_se: standard error of the effect size from the joint analysis; pJ: p-value from the joint analysis; LD_r: LD correlation between the SNP i and SNP i + 1 for the SNPs on the list; rank: rank of the independent association signal.

**Table S7. *Cis*- and *trans*-sGene overlap with GTEx (whole blood).** The results from the *cis*- and *trans*-sQTL analyses for a total of 6,881 genes were compared to the results from the GTEx v8 in whole blood tissue. Headers are: sGene: gene ID (Ensembl v99); INTERVAL_*cis*-sQTL: TRUE if the gene has a *cis*-sQTL, FALSE otherwise; INTERVAL_*trans*-sQTL: TRUE if the gene has a *trans*-sQTL; GTEX_*cis*-sQTL: TRUE if a *cis*-sQTL is also found in GTEx for the gene; GTEX_*trans*-sQTL: TRUE if a *trans*-sQTL is also found in GTEx for the gene.

**Table S8. Colocalization results between *cis*-eQTL and *cis*-sQTL.** Colocalized results (PP3+PP4>09 and PP4/PP3>3) from the colocalization analysis to identify the presence of a shared causal variant between *cis*-eQTL and *cis*-sQTL. Headers are: chromosome: chromosome; splice: splicing event ID (chr:start:end:cluster_id_strand) for the *cis*-sQTL; gene: gene ID (Ensembl v99) for the *cis*-eQTL; gene_symbol: gene name (Ensembl v99); SNP_splice_rank: rank of the lead SNP after conditional analysis for the *cis*-sQTL (0 means that no conditional analysis was performed); SNP_splice: variant ID of the lead SNP after conditional analysis for the *cis*-sQTL (“unconditioned” means no conditional analysis was performed); SNP_gene_rank: rank of the lead SNP after conditional analysis for the *cis*-eQTL (0 means that no conditional analysis was performed); SNP_gene: variant id of the lead SNP after conditional analysis for the *cis*-eQTL (“unconditioned” means no conditional analysis was performed); H0,H1,H2,H3,H4 are posterior probabilities of the hypotheses 0, 1, 2, 3 and 4.

**Table S9. *Trans*-eQTLs results.** This table contains all significant 8,834 SNP-gene associations for the *trans*-eQTL analysis. Headers are: phenotype_id: gene ID (Ensembl v99); gene_name: gene name (Ensembl v99); chrpheno: chromosome location of the gene; variant_id: variant ID; chrgeno: chromosome location of the variant; pos_b38: genomic position of the variant in GRCh38; pos_b37: genomic position of the variant in GRCh37; effect_allele: effect allele; other_allele: other allele; af: allele frequency of the effect allele; chrpheno: chromosome location of the gene; gene_name: gene name from Ensembl; b: effect size of the effect allele; b_se: standard error for the effect size estimate; pval: p-value.

**Table S10. Enriched GO terms in e/sQTL results.** From the enrichment analysis, 32 GO terms were found enriched for *cis*-eGenes with a *trans*-association with a distant gene expression, and 10 GO terms were found enriched for a *trans*-association with a distant splicing event. Headers are: Gene set: description of the gene set for enrichment; Gene background: description of the gene list background for enrichment; GO term ID: GO term ID; GO term name: GO term name; category: category of the GO term (biological process, molecular function or cell component); p-value: p-value of the Fisher exact test; FDR p-value: p-value after FDR multiple testing correction.

**Table S11. *Trans*-sQTLs results.** This table contains all significant 4,093 SNP-splicing event associations for the *trans*-sQTL analysis. Headers are: phenotype_id: splicing event ID (chr:start:end:cluster_id_strand); gene_id: gene ID (Ensembl v99); gene_name: gene name (Ensembl v99); chrpheno: chromosome location of the gene; variant_id: variant ID; chrgeno: chromosome location of the variant; pos_b38: genomic position of the variant in GRCh38; pos_b37: genomic position of the variant in GRCh37; effect_allele: effect allele; other_allele: other allele; af: allele frequency of the effect allele; chrpheno: chromosome location of the gene; gene_name: gene name from Ensembl; b: effect size of the effect allele; b_se: standard error for the effect size estimate; pval: p-value.

**Table S12. Summary of colocalization results between *cis*-eQTL or -sQTL with molecular traits.**

**Table S13. Colocalized results between *cis*-eGenes and molecular traits.** This table contains all the colocalized results (PP3+PP4>09 and PP4/PP3>3) from colocalization analyses between gene expression and proteomic and metabolomic molecular traits from INTERVAL cohort and external studies. Headers are: Study: name of the study cohort(s) from which the molecular trait QTL summary statistics were obtained; Technology: short description of the technology or platform being used to measure the molecular trait; chromosome: chromosome location of the *cis*-eQTL; gene: gene Ensembl ID; gene_symbol: gene name from Ensembl; phen: molecular phenotype ID (see Table S15 for more information); SNP_gene_rank: rank of the lead SNP after conditional analysis for the *cis*-eQTL (0 means that no conditional analysis was performed); SNP_gene: variant id of the lead SNP after conditional analysis for the *cis*-eQTL (“unconditioned” means no conditional analysis was performed); SNP_phen_rank: rank of the lead SNP after conditional analysis for the molecular trait (0 means that no conditional analysis was performed); SNP_phen: variant ID of the lead SNP after conditional analysis for the molecular trait (“unconditioned” means no conditional analysis was performed); H0, H1, H2, H3 and H4 are posterior probabilities of the hypotheses 0, 1, 2, 3 and 4.

**Table S14. Colocalized results between *cis*-sQTLs and molecular traits.** This table contains all the colocalized results (PP3+PP4>09 and PP4/PP3>3) from colocalization analyses between splice intron usage and proteomic and metabolomic molecular traits from INTERVAL cohort and external studies. Headers are: Study: name of the study cohort(s) from which the molecular trait QTL summary statistics were obtained; Technology: short description of the technology or platform being used to measure the molecular trait; chromosome: chromosome location of the *cis*-sQTL; splice: splicing event ID; gene: mapped gene Ensembl IDs; gene_symbol: mapped gene names; phen: molecular phenotype ID (see Table S15 for more information); SNP_splice_rank: rank of the lead SNP after conditional analysis for the *cis*-sQTL (0 means that no conditional analysis was performed); SNP_splice: variant ID of the lead SNP after conditional analysis for the *cis*-sQTL (“unconditioned” means no conditional analysis was performed); SNP_phen_rank: rank of the lead SNP after conditional analysis for the molecular trait (0 means that no conditional analysis was performed); SNP_phen: variant ID of the lead SNP after conditional analysis for the molecular trait (“unconditioned” means no conditional analysis was performed); H0, H1, H2, H3 and H4 are posterior probabilities of the hypotheses 0, 1, 2, 3 and 4.

**Table S15. Summary of colocalized results for *cis*-eGenes and *cis*-sGenes.** This table contains the summarised colocalization result for all *cis*-e/sGenes and molecular traits being analysed. Headers are : Study: name of the study cohort(s) from which the molecular trait QTL summary statistics were obtained; Technology: short description of the technology or platform being used to measure the molecular trait; chromosome: chromosome location of the *cis*-e/sQTL; gene: gene Ensembl ID; gene_symbol: gene name; phen_id: molecular phenotype ID; phen_desc: description of the molecular phenotype; colocalized_eQTL: TRUE if there is a colocalized signal (PP3+PP4>09 and PP4/PP3>3) with a *cis*-eQTL, FALSE otherwise; colocalized_sQTL: TRUE if there is a colocalized signal (PP3+PP4>09 and PP4/PP3>3) with a *cis*-sQTL, FALSE otherwise; RI: retention time/index information for Metabolon-measured traits; MASS: mass (g/mol) information for Metabolon-measured traits.

**Table S16. pQTL colocalizing with a *cis*-sQTL for the excision of a transmembrane domain in the encoding mRNA.** Colocalization results (PP3+PP4>09 and PP4/PP3>3) between proteins and splicing events excising a transmembrane domain in the encoding mRNA. Headers are: Study: name of the study cohort(s) from which the molecular trait QTL summary statistics were obtained; Technology: short description of the technology or platform being used to measure the molecular trait; chromosome: chromosome location of the *cis*-sQTL; splice: splicing event ID; gene: mapped gene Ensembl IDs; gene_symbol: mapped gene names; phen: molecular phenotype ID (see Table S15 for more information); SNP_splice_rank: rank of the lead SNP after conditional analysis for the *cis*-sQTL (0 means that no conditional analysis was performed); SNP_splice: variant ID of the lead SNP after conditional analysis for the *cis*-sQTL (“unconditioned” means no conditional analysis was performed); SNP_phen_rank: rank of the lead SNP after conditional analysis for the molecular trait (0 means that no conditional analysis was performed); SNP_phen: variant ID of the lead SNP after conditional analysis for the molecular trait (“unconditioned” means no conditional analysis was performed); H0, H1, H2, H3 and H4 are posterior probabilities of the hypotheses 0, 1, 2, 3 and 4.

**Table S17. Mediation results with gene expression as mediator.** This table contains all results from mediation analysis with gene expression as mediator (a total of 15,114 mediation models). Headers are: Technology: short description of the technology or platform being used to measure the molecular trait; chromosome: chromosome location of the *cis*-eQTL; gene; gene Ensembl ID; gene_name: gene name; gene_biotype: gene biotype from Ensembl; phen_id: molecular phenotype ID; variant: variant ID; nsamples: number of samples for the mediation analysis; correlation: Pearson correlation coefficient between the gene expression and the molecular phenotype; DE_est: direct effect estimate; DE_se: standard error of the direct effect estimate; DE_z: direct effect Z-score; DE_p: direct effect p-value; ME_est: mediated effect estimate; ME_se: standard error of the mediated effect estimate; ME_z: mediated effect Z-score; ME_p: mediated effect p-value; TE_est: total effect estimate; TE_se: standard error of the total effect estimate; TE_z: total effect Z-score; TE_p: total effect p-value; prop_med_est: proportion of the mediated effect over the total effect; Bonferroni_threshold: significance threshold after Bonferroni multiple testing correction; ME_significant: TRUE if the mediated effect is significant, FALSE otherwise.

**Table S18. Mediation results with splicing as mediator.** This table contains all results from mediation analysis with splicing as mediator (a total of 11,445 mediation models). Headers are: Technology: short description of the technology or platform being used to measure the molecular trait; chromosome: chromosome location of the *cis*-sQTL; splice: splicing event ID; gene; mapped gene Ensembl IDs; gene_name: mapped gene names; phen_id: molecular phenotype ID; variant: variant ID; nsamples: number of samples for the mediation analysis; DE_est: direct effect estimate; DE_se: standard error of the direct effect estimate; DE_z: direct effect Z-score; DE_p: direct effect p-value; ME_est: mediated effect estimate; ME_se: standard error of the mediated effect estimate; ME_z: mediated effect Z-score; ME_p: mediated effect p-value; TE_est: total effect estimate; TE_se: standard error of the total effect estimate; TE_z: total effect Z-score; TE_p: total effect p-value; prop_med_est: proportion of the mediated effect over the total effect; Bonferroni_threshold: significance threshold after Bonferroni multiple testing correction; ME_significant: TRUE if the mediated effect is significant, FALSE otherwise.

**Table S19. List of FinnGen traits being analyzed.** This table contains the list of 20 phenotypes from FinnGen. Headers are: phenocode: short name of the phenotype; name: long name of the phenotype; category: phenotype category; num_cases: number of cases; num_controls: number of controls; path_https: https link for data access.

**Table S20. Colocalization results between *cis*-eQTLs and FinnGen traits.** This table contains all colocalized results (PP3+PP4>09 and PP4/PP3>3) from colocalization analyses between gene expression and 20 FinnGen traits. Headers are: chromosome: chromosome location of the *cis*-eQTL; gene: gene Ensembl ID; gene_symbol: gene name from Ensembl; phen: FinnGen phenotype; SNP_gene_rank: rank of the lead SNP after conditional analysis for the *cis*-eQTL (0 means that no conditional analysis was performed); SNP_gene: variant id of the lead SNP after conditional analysis for the *cis*-eQTL (“unconditioned” means no conditional analysis was performed); SNP_phen_rank: rank of the lead SNP after conditional analysis for the FinnGen phenotype (0 means that no conditional analysis was performed); SNP_phen: variant ID of the lead SNP after conditional analysis for the FinnGen phenotype (“unconditioned” means no conditional analysis was performed); H0, H1, H2, H3 and H4 are posterior probabilities of the hypotheses 0, 1, 2, 3 and 4.

**Table S21. Colocalization results between *cis*-sQTLs and FinnGen traits.** This table contains all the colocalized results (PP3+PP4>09 and PP4/PP3>3) from colocalization analyses between splice intron usage and 20 FinnGen traits. Headers are: chromosome: chromosome location of the *cis*-sQTL; splice: splicing event ID; gene: mapped gene Ensembl ID; gene_symbol: mapped gene name; phen: FinnGen phenotype; SNP_splice_rank: rank of the lead SNP after conditional analysis for the *cis*-sQTL (0 means that no conditional analysis was performed); SNP_splice: variant ID of the lead SNP after conditional analysis for the *cis*-sQTL (“unconditioned” means no conditional analysis was performed); SNP_phen_rank: rank of the lead SNP after conditional analysis for the FinnGen phenotype (0 means that no conditional analysis was performed); SNP_phen: variant ID of the lead SNP after conditional analysis for the FinnGen phenotype (“unconditioned” means no conditional analysis was performed); H0, H1, H2, H3 and H4 are posterior probabilities of the hypotheses 0, 1, 2, 3 and 4.

**Table S22. Colocalization results between *cis*-eQTLs and COVID-19 loci.** This table contains all colocalized results (PP3+PP4>09 and PP4/PP3>3) from colocalization analyses between gene expression and 4 COVID-19 phenotypes. Headers are: chromosome: chromosome location of the *cis*-eQTL; gene: gene Ensembl ID; gene_symbol: gene name from Ensembl; phen: COVID-19 phenotype (A2, very severe respiratory confirmed COVID-19 patients versus population; B1, hospitalized versus non-hospitalized COVID-19 patients; and B2, hospitalized COVID-19 patients versus population); SNP_gene_rank: rank of the lead SNP after conditional analysis for the *cis*-eQTL (0 means that no conditional analysis was performed); SNP_gene: variant id of the lead SNP after conditional analysis for the *cis*-eQTL (“unconditioned” means no conditional analysis was performed); SNP_phen_rank: rank of the lead SNP after conditional analysis for the COVID-19 phenotype (0 means that no conditional analysis was performed); SNP_phen: variant ID of the lead SNP after conditional analysis for the COVID-19 phenotype (“unconditioned” means no conditional analysis was performed); H0, H1, H2, H3 and H4 are posterior probabilities of the hypotheses 0, 1, 2, 3 and 4.

**Table S23. Colocalization results between *cis*-sQTLs and COVID-19 loci.** This table contains all the colocalized results (PP3+PP4>09 and PP4/PP3>3) from colocalization analyses between splice intron usage and 4 COVID-19 phenotypes. Headers are: chromosome: chromosome location of the *cis*-sQTL; splice: splicing event ID; gene: mapped gene Ensembl ID; gene_symbol: mapped gene name; phen: COVID-19 phenotype (A2, very severe respiratory confirmed COVID-19 patients versus population; B1, hospitalized versus non-hospitalized COVID-19 patients; and B2, hospitalized COVID-19 patients versus population); SNP_splice_rank: rank of the lead SNP after conditional analysis for the *cis*-sQTL (0 means that no conditional analysis was performed); SNP_splice: variant ID of the lead SNP after conditional analysis for the *cis*-sQTL (“unconditioned” means no conditional analysis was performed); SNP_phen_rank: rank of the lead SNP after conditional analysis for the COVID-19 phenotype (0 means that no conditional analysis was performed); SNP_phen: variant ID of the lead SNP after conditional analysis for the COVID-19 phenotype (“unconditioned” means no conditional analysis was performed); H0, H1, H2, H3 and H4 are posterior probabilities of the hypotheses 0, 1, 2, 3 and 4.

**Table S24. List of included covariates for the different analyses.** This table contains the list of all covariates for PEER factor computation, eQTL and sQTL analyses.

**Table S25. Covariate description in the INTERVAL cohort with RNA-seq data.** For quantitative covariates, the columns “Mean”, “Median”, “Min”, “Max” are the mean, median, minimum and maximum values as distribution descriptors. For categorical covariate, “Categories information” is a short description of the variable modalities.

**Table S26. List of summary statistics for colocalization analyses.** Headers are: PMID: Pubmed ID; Trait category: indicates if the trait is a proteomic or metabolomic molecular trait or a disease trait; Technology: short description of the technology or platform being used to measure the molecular trait; Population study: cohort names for each study; Sample size for QTL analysis/GWAS: sample sizes for each study; Number of features: number of selected analyzed traits for each study; Sample overlap with RNA-seq data for mediation analysis: number of individuals overlapping with the RNA-seq if a mediation analysis was also conducted in addition to the colocalization analysis (only available for INTERVAL molecular traits); Data availability: link to the publicly available summary statistics, or mode of obtention if non publicly available.

## References

1. Loos, R.J.F. 15 years of genome-wide association studies and no signs of slowing down. Nat Commun 11, 5900 (2020).

2. Albert, F.W. & Kruglyak, L. The role of regulatory variation in complex traits and disease. Nat Rev Genet 16, 197–212 (2015).

3. Neumeyer, S., Hemani, G. & Zeggini, E. Strengthening Causal Inference for Complex Disease Using Molecular Quantitative Trait Loci. Trends Mol Med 26, 232–241 (2020).

4. Suhre, K. et al. Human metabolic individuality in biomedical and pharmaceutical research. Nature 477, 54–60 (2011).

5. Sun, B.B. et al. Genomic atlas of the human plasma proteome. Nature 558, 73–79 (2018).

6. Suhre, K., McCarthy, M.I. & Schwenk, J.M. Genetics meets proteomics: perspectives for large population-based studies. Nat Rev Genet 22, 19–37 (2021).

7. Kim-Hellmuth, S. et al. Cell type-specific genetic regulation of gene expression across human tissues. Science 369(2020).

8. Ferkingstad, E. et al. Large-scale integration of the plasma proteome with genetics and disease. Nat Genet 53, 1712–1721 (2021).

9. Vosa, U. et al. Large-scale cis- and trans-eQTL analyses identify thousands of genetic loci and polygenic scores that regulate blood gene expression. Nat Genet 53, 1300–1310 (2021).

10. Surendran, P. et al. Rare and common genetic determinants of metabolic individuality and their effects on human health. Nat Med 28, 2321–2332 (2022).

11. Julkunen, H. et al. Atlas of plasma NMR biomarkers for health and disease in 118,461 individuals from the UK Biobank. Nat Commun 14, 604 (2023).

12. Sun, B.B. et al. Plasma proteomic associations with genetics and health in the UK Biobank. Nature 622, 329–338 (2023).

13. Suhre, K. et al. Connecting genetic risk to disease end points through the human blood plasma proteome. Nat Commun 8, 14357 (2017).

14. Burgess, S. et al. Guidelines for performing Mendelian randomization investigations: update for summer 2023. Wellcome Open Res 4, 186 (2019).

15. Moore, C. et al. The INTERVAL trial to determine whether intervals between blood donations can be safely and acceptably decreased to optimise blood supply: study protocol for a randomised controlled trial. Trials 15, 363 (2014).

16. Di Angelantonio, E. et al. Efficiency and safety of varying the frequency of whole blood donation (INTERVAL): a randomised trial of 45 000 donors. Lancet 390, 2360–2371 (2017).

17. Chen, L. et al. Systematic Mendelian randomization using the human plasma proteome to discover potential therapeutic targets for stroke. Nat Commun 13, 6143 (2022).

18. SCALLOP Consortium. Mapping pQTLs of circulating inflammatory proteins identifies drivers of immune-related disease risk and novel therapeutic targets. medRxiv (2023).

19. Riveros-Mckay, F. et al. The influence of rare variants in circulating metabolic biomarkers. PLoS Genet 16, e1008605 (2020).

20. Karjalainen, M.K. et al. Genome-wide characterization of circulating metabolic biomarkers reveals substantial pleiotropy and novel disease pathways. medRxiv (2023).

21. Wang, H. et al. Long noncoding RNA FAM157C contributes to clonal proliferation in paroxysmal nocturnal hemoglobinuria. Ann Hematol 102, 299–309 (2023).

22. Yamaguchi, K. et al. Splicing QTL analysis focusing on coding sequences reveals mechanisms for disease susceptibility loci. Nat Commun 13, 4659 (2022).

23. GTEx Consortium. The GTEx Consortium atlas of genetic regulatory effects across human tissues. Science 369, 1318–1330 (2020).

24. Lambert, S.A. et al. The Human Transcription Factors. Cell 172, 650–665 (2018).

25. Keyvani Chahi, A., et al. PLAG1 dampens protein synthesis to promote human hematopoietic stem cell self-renewal. Blood 140, 992–1008 (2022).

26. Pietzner, M. et al. Mapping the proteo-genomic convergence of human diseases. Science 374, eabj1541 (2021).

27. Xing, Y., Xu, Q. & Lee, C. Widespread production of novel soluble protein isoforms by alternative splicing removal of transmembrane anchoring domains. FEBS Lett 555, 572–8 (2003).

28. Paronetto, M.P., Passacantilli, I. & Sette, C. Alternative splicing and cell survival: from tissue homeostasis to disease. Cell Death Differ 23, 1919–1929 (2016).

29. Goodwin, R.G. et al. Cloning of the human and murine interleukin-7 receptors: demonstration of a soluble form and homology to a new receptor superfamily. Cell 60, 941–51 (1990).

30. Lust, J.A. et al. Isolation of an mRNA encoding a soluble form of the human interleukin-6 receptor. Cytokine 4, 96–100 (1992).

31. Briso, E.M., Dienz, O. & Rincon, M. Cutting edge: soluble IL-6R is produced by IL-6R ectodomain shedding in activated CD4 T cells. J Immunol 180, 7102–6 (2008).

32. Garbers, C. et al. The interleukin-6 receptor Asp358Ala single nucleotide polymorphism rs2228145 confers increased proteolytic conversion rates by ADAM proteases. Biochim Biophys Acta 1842, 1485–94 (2014).

33. Hormozdiari, F. et al. Colocalization of GWAS and eQTL Signals Detects Target Genes. Am J Hum Genet 99, 1245–1260 (2016).

34. Kurki, M.I. et al. Author Correction: FinnGen provides genetic insights from a well-phenotyped isolated population. Nature 615, E19 (2023).

35. Akbari, P. et al. A genome-wide association study of blood cell morphology identifies cellular proteins implicated in disease aetiology. Nat Commun 14, 5023 (2023).

36. Gregory, S.G. et al. Interleukin 7 receptor alpha chain (IL7R) shows allelic and functional association with multiple sclerosis. Nat Genet 39, 1083–91 (2007).

37. Ahn, Y.H. et al. Secreted tryptophanyl-tRNA synthetase as a primary defence system against infection. Nat Microbiol 2, 16191 (2016).

38. Gioelli, N. et al. Neuropilin 1 and its inhibitory ligand mini-tryptophanyl-tRNA synthetase inversely regulate VE-cadherin turnover and vascular permeability. Nat Commun 13, 4188 (2022).

39. Covid-19 Host Genetics Initiative. A first update on mapping the human genetic architecture of COVID-19. Nature 608, E1–E10 (2022).

40. Bonnevie-Nielsen, V. et al. Variation in antiviral 2’,5’-oligoadenylate synthetase (2’5’AS) enzyme activity is controlled by a single-nucleotide polymorphism at a splice-acceptor site in the OAS1 gene. Am J Hum Genet 76, 623–33 (2005).

41. Huffman, J.E. et al. Multi-ancestry fine mapping implicates OAS1 splicing in risk of severe COVID-19. Nat Genet 54, 125–127 (2022).

42. Smieszek, S.P., Polymeropoulos, V.M., Xiao, C., Polymeropoulos, C.M. & Polymeropoulos, M.H. Loss-of-function mutations in IFNAR2 in COVID-19 severe infection susceptibility. J Glob Antimicrob Resist 26, 239–240 (2021).

43. Novick, D., Cohen, B., Tal, N. & Rubinstein, M. Soluble and membrane-anchored forms of the human IFN-alpha/beta receptor. J Leukoc Biol 57, 712–8 (1995).

44. Yaugel-Novoa, M., Bourlet, T., Longet, S., Botelho-Nevers, E. & Paul, S. Association of IFNAR1 and IFNAR2 with COVID-19 severity. Lancet Microbe 4, e487 (2023).

45. Finucane, H.K. et al. Partitioning heritability by functional annotation using genome-wide association summary statistics. Nat Genet 47, 1228–35 (2015).

46. Xu, Y. et al. An atlas of genetic scores to predict multi-omic traits. Nature 616, 123–131 (2023).

47. Li, Y.I. et al. RNA splicing is a primary link between genetic variation and disease. Science 352, 600–4 (2016).

48. Ritchie, S.C. et al. Quality control and removal of technical variation of NMR metabolic biomarker data in ∼120,000 UK Biobank participants. Sci Data 10, 64 (2023).

49. Liu, X. et al. Functional Architectures of Local and Distal Regulation of Gene Expression in Multiple Human Tissues. Am J Hum Genet 100, 605–616 (2017).

50. Qi, T. et al. Identifying gene targets for brain-related traits using transcriptomic and methylomic data from blood. Nat Commun 9, 2282 (2018).

51. van der Wijst, M. et al. The single-cell eQTLGen consortium. Elife 9(2020).

52. Cuomo, A.S.E., Nathan, A., Raychaudhuri, S., MacArthur, D.G. & Powell, J.E. Single-cell genomics meets human genetics. Nat Rev Genet 24, 535–549 (2023).

53. Fort, A. et al. MBV: a method to solve sample mislabeling and detect technical bias in large combined genotype and sequencing assay datasets. Bioinformatics 33, 1895–1897 (2017).

54. Dobin, A. et al. STAR: ultrafast universal RNA-seq aligner. Bioinformatics 29, 15–21 (2013).

55. Liao, Y., Smyth, G.K. & Shi, W. featureCounts: an efficient general purpose program for assigning sequence reads to genomic features. Bioinformatics 30, 923–30 (2014).

56. Chang, C.C. et al. Second-generation PLINK: rising to the challenge of larger and richer datasets. Gigascience 4, 7 (2015).

57. Robinson, M.D. & Oshlack, A. A scaling normalization method for differential expression analysis of RNA-seq data. Genome Biol 11, R25 (2010).

58. Aulchenko, Y.S., Ripke, S., Isaacs, A. & van Duijn, C.M. GenABEL: an R library for genome-wide association analysis. Bioinformatics 23, 1294–6 (2007).

59. Cotto, K.C. et al. Integrated analysis of genomic and transcriptomic data for the discovery of splice-associated variants in cancer. Nat Commun 14, 1589 (2023).

60. Li, Y.I. et al. Annotation-free quantification of RNA splicing using LeafCutter. Nat Genet 50, 151–158 (2018).

61. Astle, W.J. et al. The Allelic Landscape of Human Blood Cell Trait Variation and Links to Common Complex Disease. Cell 167, 1415–1429 e19 (2016).

62. Zhao, H. et al. CrossMap: a versatile tool for coordinate conversion between genome assemblies. Bioinformatics 30, 1006–7 (2014).

63. Stegle, O., Parts, L., Piipari, M., Winn, J. & Durbin, R. Using probabilistic estimation of expression residuals (PEER) to obtain increased power and interpretability of gene expression analyses. Nat Protoc 7, 500–7 (2012).

64. Taylor-Weiner, A. et al. Scaling computational genomics to millions of individuals with GPUs. Genome Biol 20, 228 (2019).

65. Ongen, H., Buil, A., Brown, A.A., Dermitzakis, E.T. & Delaneau, O. Fast and efficient QTL mapper for thousands of molecular phenotypes. Bioinformatics 32, 1479–85 (2016).

66. Yang, J., Lee, S.H., Goddard, M.E. & Visscher, P.M. GCTA: a tool for genome-wide complex trait analysis. Am J Hum Genet 88, 76–82 (2011).

67. Yang, J. et al. Conditional and joint multiple-SNP analysis of GWAS summary statistics identifies additional variants influencing complex traits. Nat Genet 44, 369–75, S1-3 (2012).

68. Ashburner, M. et al. Gene ontology: tool for the unification of biology. The Gene Ontology Consortium. Nat Genet 25, 25–9 (2000).

69. Giambartolomei, C. et al. Bayesian test for colocalisation between pairs of genetic association studies using summary statistics. PLoS Genet 10, e1004383 (2014).

70. Zheng, J. et al. Phenome-wide Mendelian randomization mapping the influence of the plasma proteome on complex diseases. Nat Genet 52, 1122–1131 (2020).

71. Mitchelmore, J., Grinberg, N.F., Wallace, C. & Spivakov, M. Functional effects of variation in transcription factor binding highlight long-range gene regulation by epromoters. Nucleic Acids Res 48, 2866–2879 (2020).

72. Peterson, C.B., Bogomolov, M., Benjamini, Y. & Sabatti, C. TreeQTL: hierarchical error control for eQTL findings. Bioinformatics 32, 2556–8 (2016).

73. Huang, Q.Q., Ritchie, S.C., Brozynska, M. & Inouye, M. Power, false discovery rate and Winner’s Curse in eQTL studies. Nucleic Acids Res 46, e133 (2018).

74. Steen, J., Loeys, T., Moerkerke, B., Vansteelandt, S. medflex: An R Package for Flexible Mediation Analysis using Natural Effect Models. J. Stat. Softw. 76, 1–46 (2017).

