## Supplementary figures for "Genetic determinants of blood gene expression and splicing and their contribution to molecular phenotypes and health outcomes"

**Figure S1.** **Correlation of *cis*-eQTL Z-scores between INTERVAL and eQTLGen.**
**Figure S2.** ***Cis*-eQTL and -sQTL independent signal distribution around TSS and gene boundaries.**
**Figure S3.** ***Cis*-eQTL and -sQTL colocalization analysis and lead SNP distribution around the TSS.**
**Figure S4.** **Transcription factor annotation in *cis*- and *trans*-QTL results.**
**Figure S5.** **Summary of splice event QC.**
**Figure S6.** **Identification of the optimal number of PEER factors (latent variation estimated from gene expression data) to be included as covariates in the eQTL analysis.**

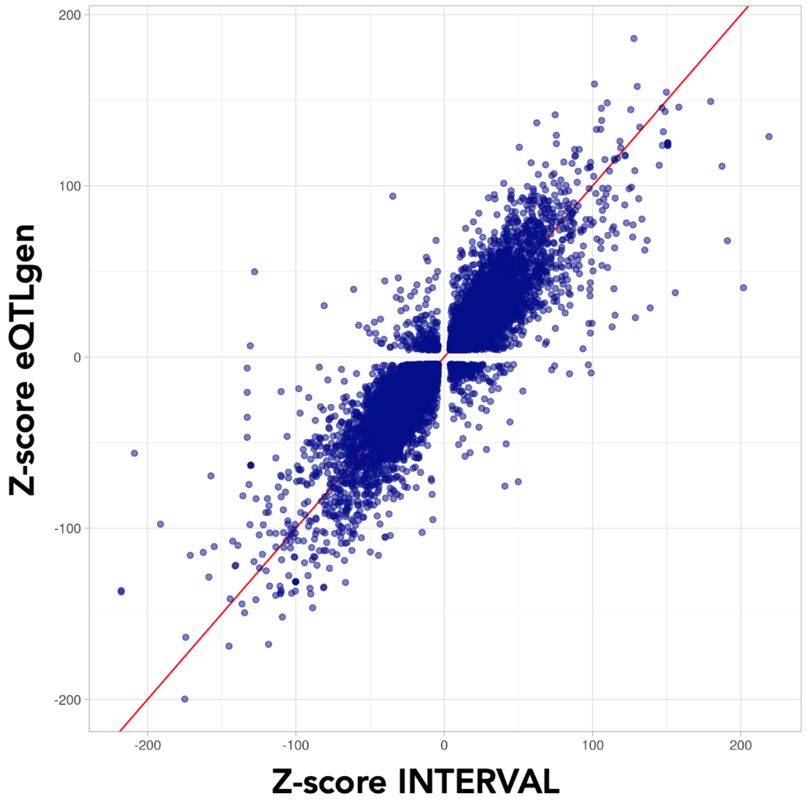


**Figure S1. Correlation of *cis*-eQTL Z-scores between INTERVAL and eQTLGen.**

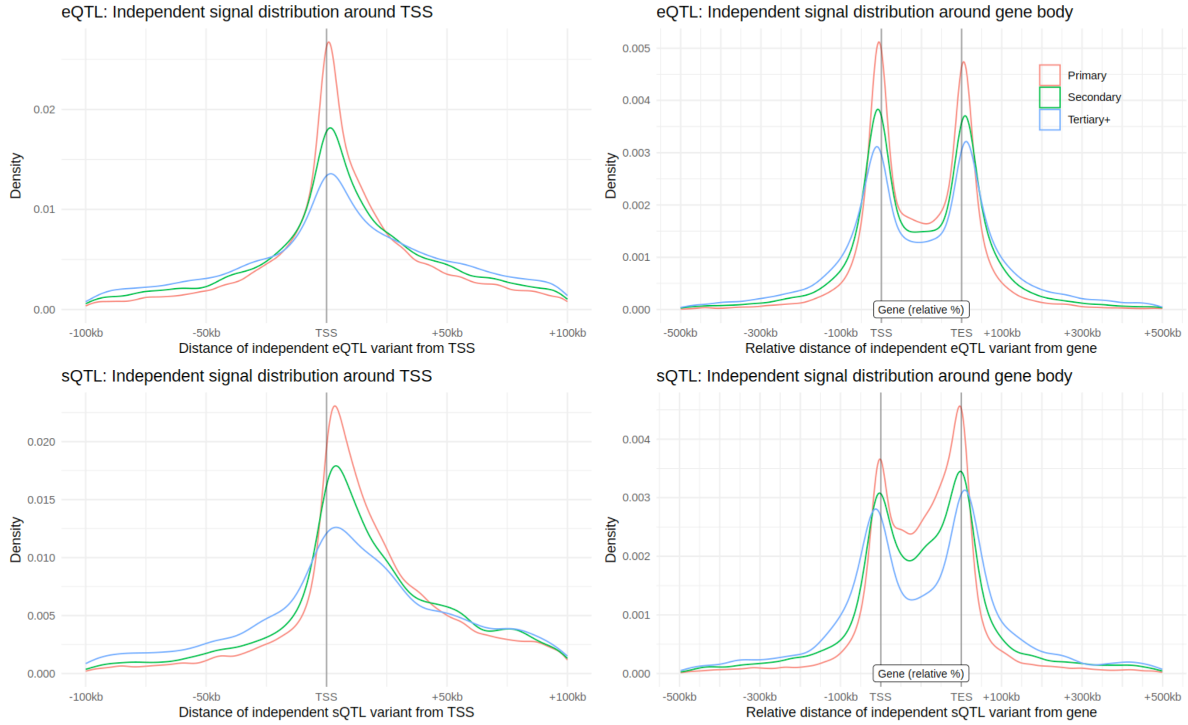


**Figure S2. *Cis*-eQTL and -sQTL independent signal distribution around TSS and gene boundaries.**

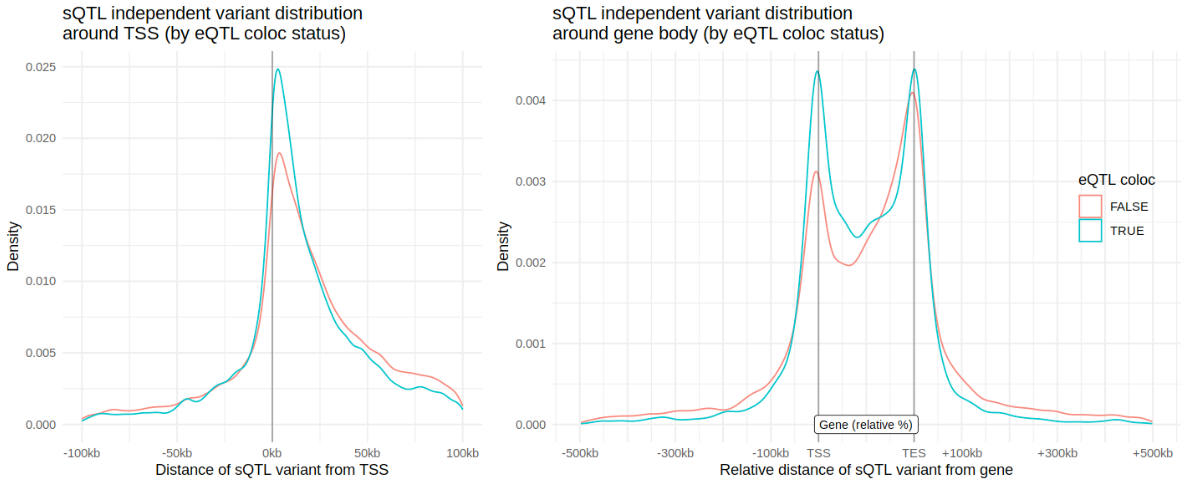


**Figure S3.** ***Cis*-eQTL and -sQTL colocalization analysis and lead SNP distribution around the TSS.**

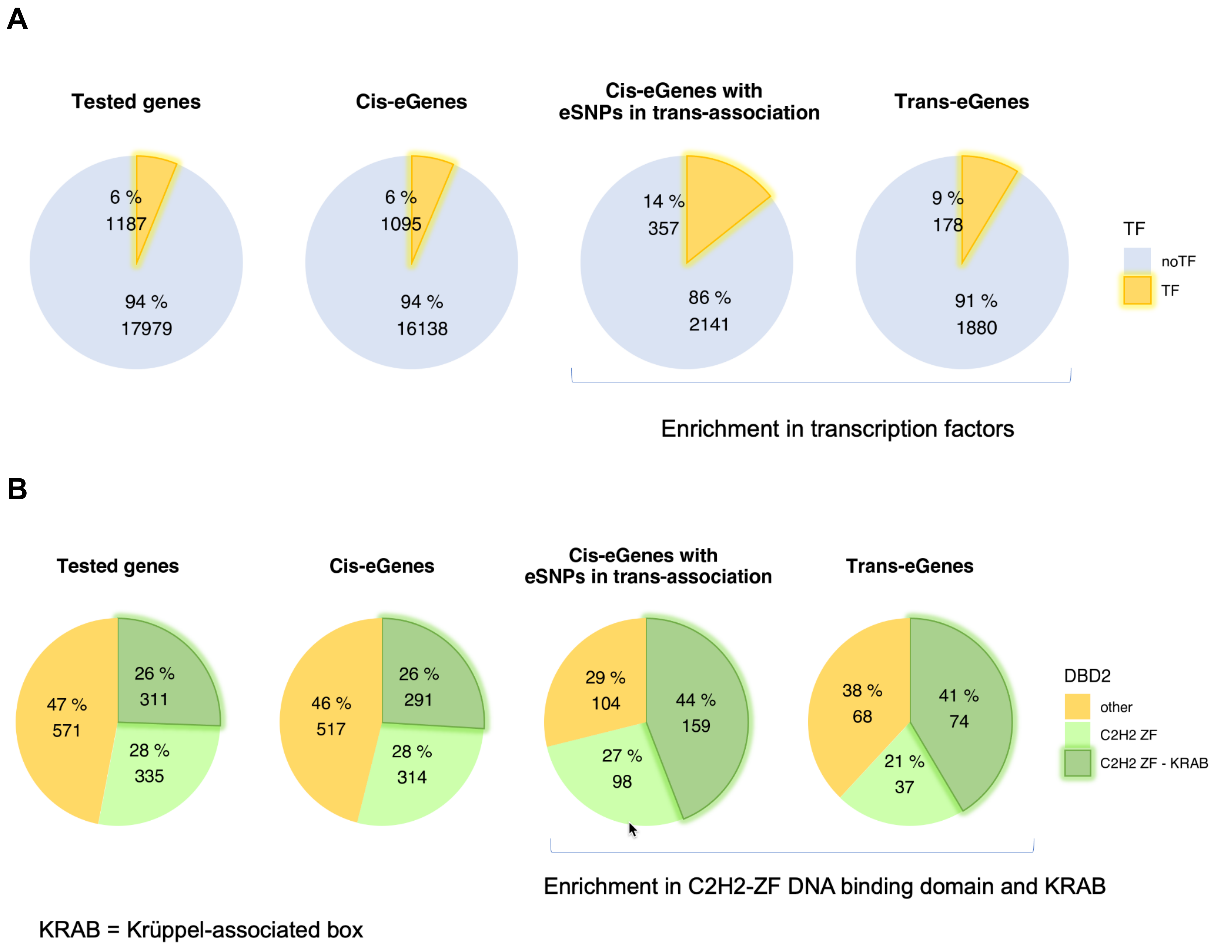


**Figure S4. Transcription factor annotation in *cis*- and *trans*-QTL results.** A. Proportion of transcription factors in *cis*- and *trans*-eGenes; B. Subcategories of transcription factors in *cis*- and *trans*-eGenes.

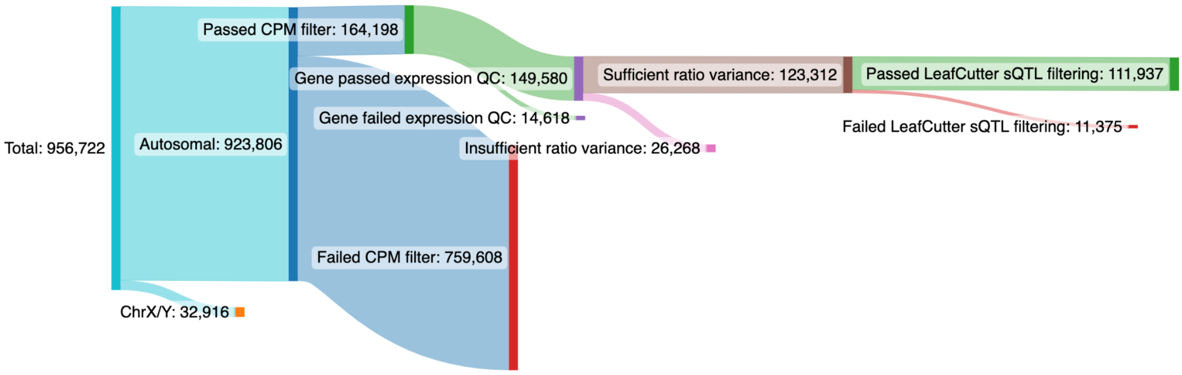


**Figure S5. Summary of splice event QC.**

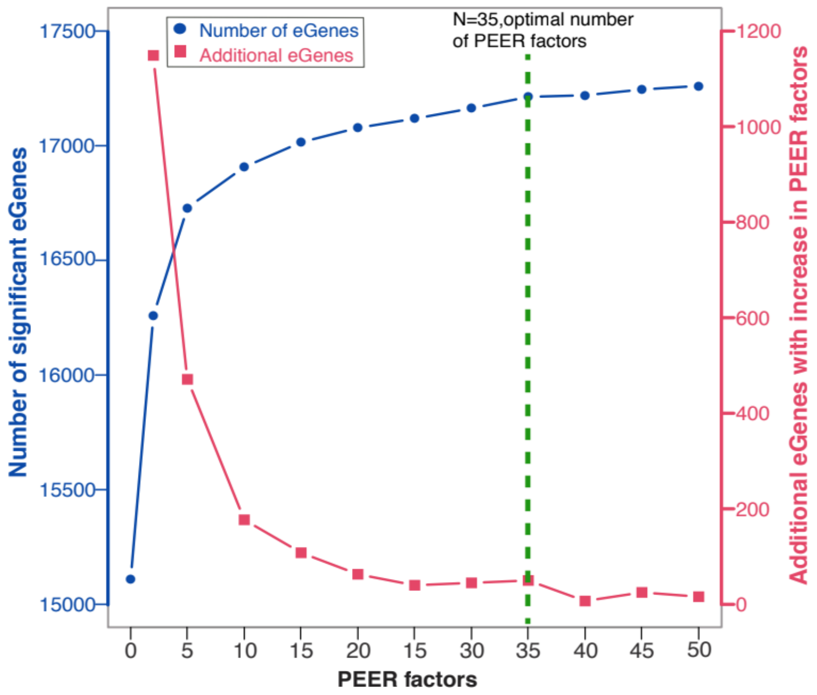


**Figure S6. Identification of the optimal number of PEER factors (latent variation estimated from gene expression data) to be included as covariates in the eQTL analysis.** PEER factors were chosen based on the maximum number of eGenes discovered (Y-axis, left) and the gain in eGenes with incremental increase in PEER factors (Y-axis, right).
